# Copper-deficiency is associated with impairments in social behavior and oligodendrocyte development via mTOR signaling pathway

**DOI:** 10.1101/2023.12.16.23300061

**Authors:** Noriyoshi Usui, Miyuki Doi, Stefano Berto, Kiwamu Matsuoka, Rio Ishida, Koichiro Irie, Nanako Nakama, Hana Miyauchi, Yuuki Fujiwara, Takahira Yamauchi, Takaharu Hirai, Michihiro Toritsuka, Min-Jue Xie, Yoshinori Kayashima, Naoko Umeda, Keiko Iwata, Kazuki Okumura, Taeko Harada, Takeshi Yoshimura, Taiichi Katayama, Masatsugu Tsujii, Hideo Matsuzaki, Manabu Makinodan, Shoichi Shimada

## Abstract

Autism spectrum disorder (ASD) is a heterogeneous disorder characterized by impaired social communication and restricted repetitive behaviors, however the biological mechanisms remain unclear. Although trace elements play essential roles in the living body, it is unclear how alterations of trace elements in ASD are involved in pathogenesis. Here we analyzed the plasma metallome and identified the alterations of 11 elements in individuals with ASD. The copper decrease was negatively correlated with ASD symptom scores. A copper-deficient mouse model reflecting the condition showed ASD-like behaviors and impaired oligodendrocyte development. In copper-deficient mice, mechanistic target of rapamycin (mTOR) signaling was reduced, and its activation by agonist improved social impairment and oligodendrocyte developmental defects. Supporting these results, white matter volumes were negatively correlated with social symptoms in individuals with ASD. Our results demonstrate that copper-deficiency contributes to ASD by causing oligodendrocytes impairment via mTOR signaling. Our findings indicate that the effects of copper-deficiency and mTOR imbalance are relevant to the pathogenesis of ASD and are potential therapeutic targets.

## INTRODUCTION

ASD is a neurodevelopmental disorder (NDD) characterized by impaired social communication, restricted repetitive behaviors and interests, and hyperesthesia/hypesthesia (*1, 2*). ASD has a strong polygenic component involving hundreds of associated genes (*3–5*). Moreover, growing evidence suggests that environmental factors, such as exposure to inflammation and harmful contaminants during pregnancy, may trigger the development of ASD through interactions with genetic factors (*6, 7*). The prevalence of ASD has been reported to be 1 in 44 (2.27%) patients in the USA (*8, 9*), however no definitive treatment has been established. Therefore, understanding the biological mechanisms underlying ASD is necessary to address those issues. We have previously reported that increases in omega-3 and omega-6 fatty acids associated with social impairment by analyzing lipidome in plasma of children with ASD (*10*). Since various elements are involved in lipid metabolism, we focused on trace elements.

The comprehensive concept of the elements that constitute the living body is called the metallome. It is well-known that the trace elements play an essential role in maintaining biological functions such as DNA and RNA synthesis, transcriptional regulation, enzyme catalysis, metabolism, redox, development, and aging (*11, 12*). Undernutrition during pregnancy causes various disease risks such as low birth weight, NDDs, and psychiatric disorders (*13, 14*). For example, zinc-deficiency during pregnancy causes fetal neural tube defects. In recent years, the trace elements in the brain function and diseases such as ASD, schizophrenia (SCZ), Alzheimer’s disease, and Parkinson’s disease have attracted attention (*15–17*), however most of the molecular mechanisms remain unknown.

The relationship between ASD and trace elements began to be mainly reported in the 2000s, and 14 elements have been reported so far, including zinc, copper, iron, magnesium, calcium, and selenium (*18–23*). However, the changes in the concentrations of single or multiple elements, and there is no consistency in age and sex of subjects, types of samples, and the results. Only a few studies have attempted to understand the pathophysiological mechanisms of ASD by focusing on elements, and most of these studies have reported only concentrations of trace elements in samples.

In this study, we conducted plasma metallomics analysis between ASD and typical developmental controls (CTL) to uncover further biological mechanisms underlying the pathophysiology of ASD. We analyzed 21 elements and showed the dyshomeostasis of metallome in individuals affected by ASD. We highlighted the decreased concentrations of plasma copper in ASD that was negative correlated with clinical scores of ASD symptoms. Copper-deficient model mice recapitulating the ASD pathology showed ASD-like social impairment, restricted and repetitive behaviors (RRBs), and developmental abnormalities of oligodendrocytes due to reduced mTOR signaling. Rescuing mTOR signaling improved the behavioral and histological phenotypes in copper-deficient mice, demonstrating that impairment of biological mechanisms caused by copper-deficiency is a one of the pathogenic factors in ASD. Our finding provides a novel perspective to understanding the biological mechanisms underlying ASD.

## RESULTS

### Metallome dyshomeostasis in individuals with ASD

In total, 112 individuals with ASD and 115 CTL were evaluated for eligibility for metallomics analysis (see Methods; fig. S1 and table S1). We conducted a comprehensive metallomics analysis targeting the trace elements in the plasma of individuals with ASD and CTL using inductively coupled plasma mass spectrometry (ICP-MS) (Fig. 1A), and quantified 21 elements (Fig. 1B to V). We further assessed the metallome dyshomeostasis by using a multivariate analysis with sex, age, and hemolysis as confounders, then found that significant decreases in the plasma concentrations of sodium, magnesium, phosphorus, sulfur, calcium, cobalt, copper, and zinc in ASD compared with CTL (Fig.1C to F, H, and K to M). We also found that significant increases in the plasma concentrations of potassium, iron, and barium in ASD (Fig. 1G, J, and U). There were no differences in the plasma concentrations of lithium, manganese, arsenic, selenium, rubidium, strontium, molybdenum, silver, cesium, and thallium (Fig. 1B, I, N to T, and V). These results indicate that plasma trace elements were altered in individuals with ASD, collectively suggesting that a dyshomeostasis in the plasma metallome is associated with pathophysiology of ASD.

**Fig. 1.**
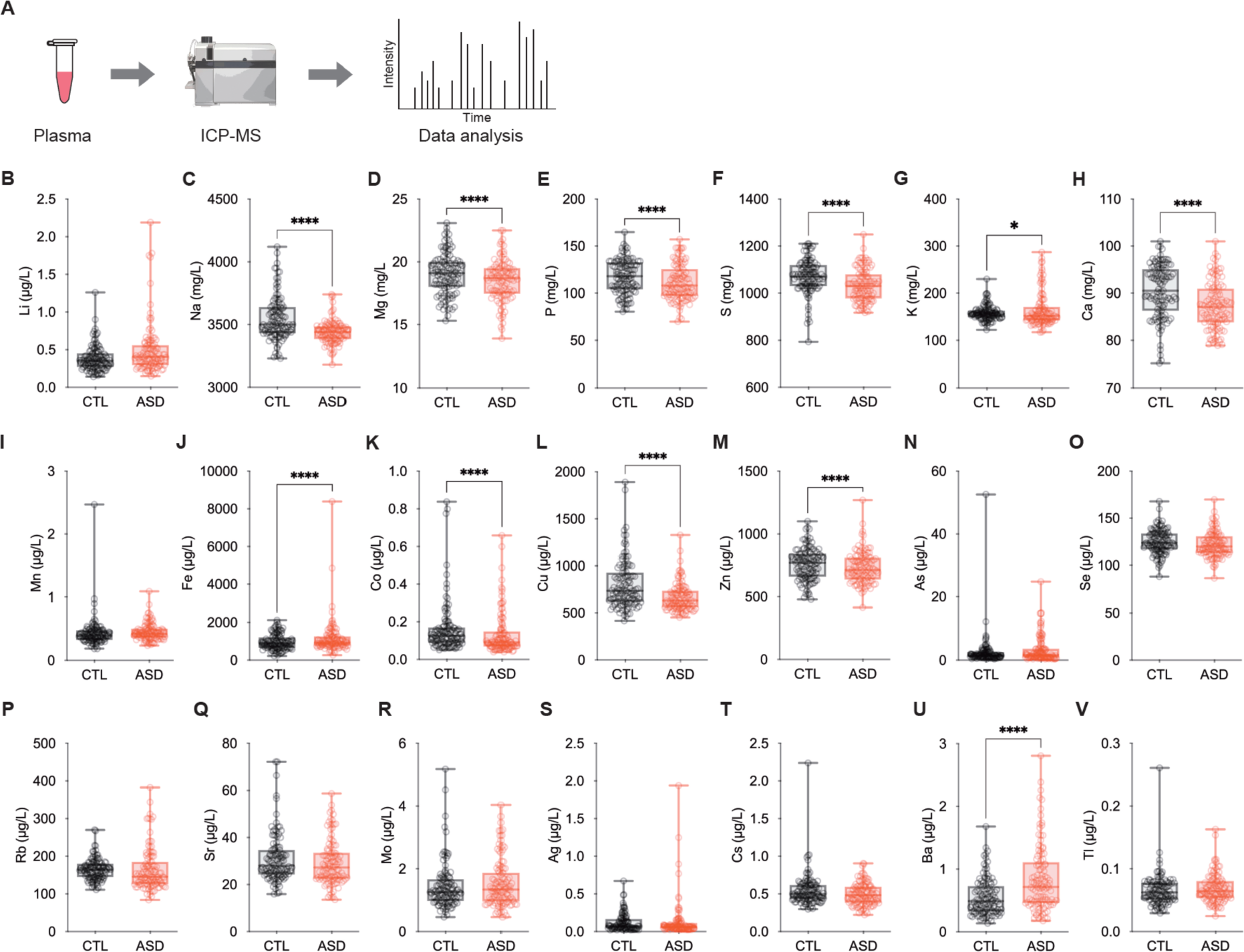
Metallomics profiling in individuals with autism spectrum disorder. (**A**) Schematic flowchart of metallomics analysis using human plasma. *ICP-MS: inductively coupled plasma mass spectrometry.* (**B** to **V**) Plasma concentrations of trace elements. *Li: lithium* (B), *Na: sodium* (C), *Mg: magnesium* (D), *P: phosphorus* (E), *S: sulfur* (F), *K: potassium* (G), *Ca: calcium* (H), *Mn: manganese* (I), *Fe: iron* (J), *Co: cobalt* (K), *Cu: copper* (L), *Zn: zinc* (M), *As: arsenic* (N), *Se: selenium* (O), *Rb: rubidium* (P), *Sr: strontium* (Q), *Mo: molybdenum* (R), *Ag: silver* (S), *Cs: cesium* (T), *Ba: barium* (U), and *Tl: thallium* (V). Significant decreases in Na, Mg, P, S, Ca, Co, Cu, and Zn concentrations were also observed in the plasma of individuals with autism spectrum disorder (ASD) compared with controls (CTL). In contrast, significant increases in K, Fe, and Ba concentrations were observed in the plasma of individuals with ASD. Data are represented as box and whiskers (minimum to maximum). Asterisks indicate ****P < 0.0001, *P < 0.05, multivariate linear regression analysis with sex, age, and hemolysis as confounders. n = 110-115/condition for Li, n = 112-115/condition for Na, Mg, P, S, K, Ca, Mn, Fe, Co, Cu, Zn, As, Se, Rb, Sr, Mo, Cs, Ba, and Tl, n = 100-107/condition for Ag.

### Plasma copper was correlated with clinical characteristics of ASD symptoms

We next evaluated whether the alterations of trace elements are associated with ASD symptoms. We performed a correlation analysis between plasma concentrations of 21 elements and clinical scores of the autism diagnostic observation schedule-second edition (ADOS-2). Interestingly, we found that only copper was negatively correlated with most of the ADOS-2 scores such as the social interaction, social affect, RRB, and total scores, except for the communication score (Fig. 2A to E).

**Fig. 2.**
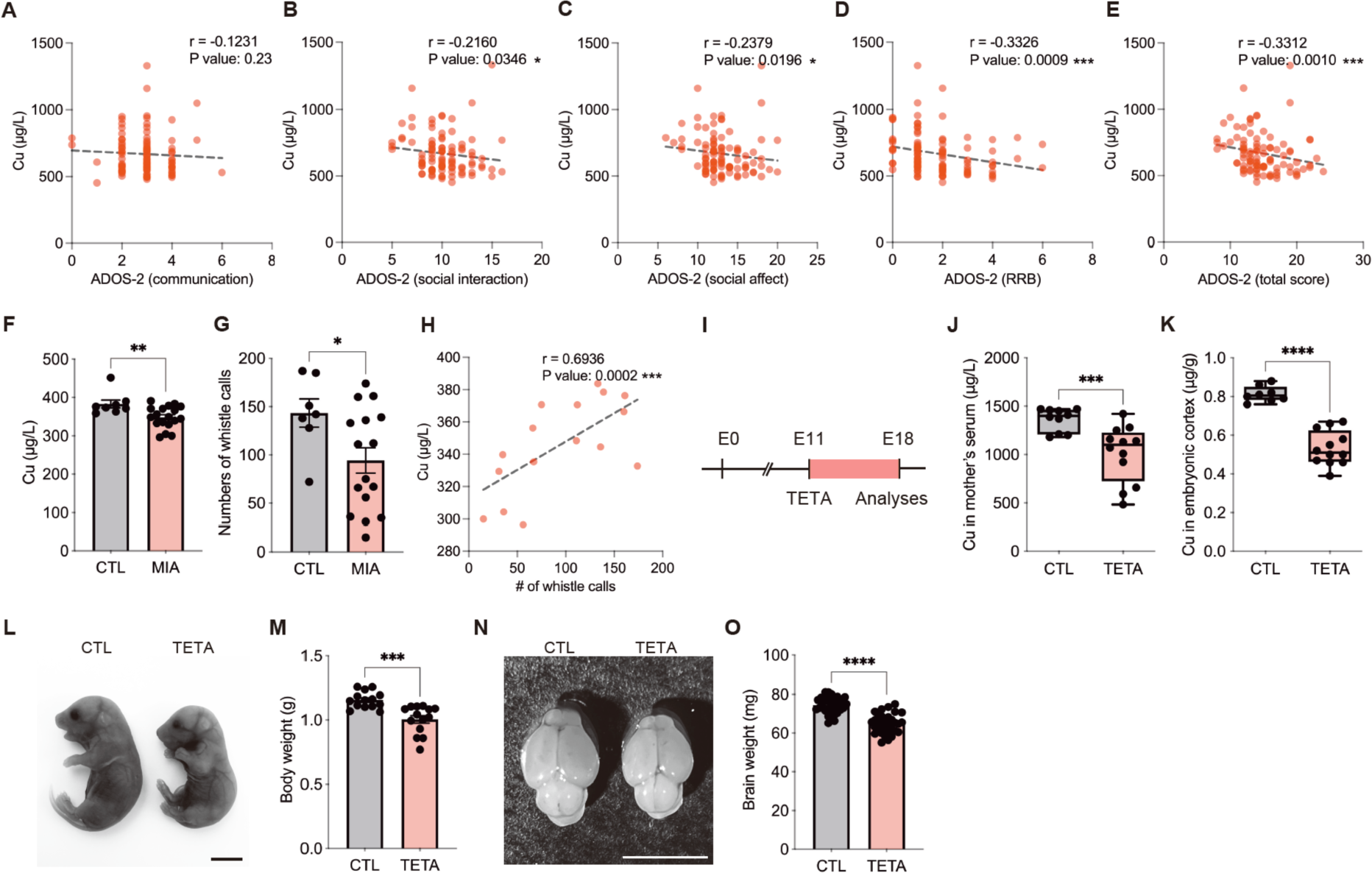
Plasma copper concentrations correlate with the characteristics of ASD symptoms. (**A** to **E**) Plasma copper concentrations correlated with clinical scores of the autism diagnostic observation schedule-second edition (ADOS-2). Plasma copper concentrations were negatively correlated with the social interaction score (B), social affect score (C), restricted and repetitive behaviors (RRB) score (D), and total score (E) of ADOS-2 in individuals with ASD, except for the communication score (A). (**F**) Quantifications of serum copper concentrations in mice at postnatal days (P) 7. Serum copper concentrations were significantly reduced in poly(I:C)-induced maternal immune activated (MIA) offspring mice compared with CTL mice. (**G**) Quantifications of mouse ultrasonic vocalizations (USVs) at P7. The numbers of USVs were significantly reduced in MIA offspring. (**H**) Correlation between serum copper concentrations and USVs in MIA offspring mice at P7. Serum copper concentrations were significantly correlated with USVs. (**I**) Experimental time course for TETA treatment to pregnant mice. (**J**) Quantifications of serum copper concentrations in mother mice at E18.5. A significant reduction of serum copper concentrations was observed in triethylenetetramine dihydrochloride-treated (TETA) mother mice. (**K**) Quantifications of cortical copper concentrations in TETA-treated embryos at E18.5. A significant reduction of cortical copper concentrations was also observed in TETA-treated embryos. (**L**) Representative images showing TETA-treated embryos show smaller bodies than CTL mice. (**M**) Quantifications of body weight at E18.5. A significant reduction of body weight was observed in TETA-treated embryos. (**N**) Representative images showing TETA-treated embryos show smaller brains than CTL mice. (**O**) Quantifications of brain weight at E18.5. A significant reduction of brain weight was also observed in TETA-treated embryos. Data are represented as means (±SEM). Asterisks indicate ****P < 0.0001, ***P < 0.001, **P < 0.01, *P < 0.05, Spearman rank correlation coefficient test, Pearson’s r correlation coefficient, unpaired t-test, Mann-Whitney test. n = 91-96/condition for correlation between copper and ADOS-2 scores, n = 8-18/condition for postnatal serum copper, n = 7-16/condition for USVs, n = 16/condition for correlation between copper and USVs, n = 10-15/condition for mother’s serum copper, n = 10-12/condition for embryonic cortical copper, n = 14/condition for embryonic body weight, n = 32-34/condition for embryonic brain weight. Scale bars: 500 μm in L, N.

To confirm the human metallome phenotype in ASD animal model, we used a polyinosinic-polycytidylic acid (poly[I:C])-induced maternal immune activation (MIA) model commonly used in ASD studies. In this MIA offspring mice, we found that reductions of serum copper concentrations and the numbers of mouse ultrasonic vocalizations (USVs) as an indicator of mouse social communication at postnatal days (P) 7, compared with CTL mice respectively (Fig. 2F and G). Interestingly, serum copper concentrations positively correlated with the numbers of USVs consistent with ASD individuals (Fig. 2H). Collectively, these results demonstrate that a reduction of copper levels is associated with ASD-like mouse phenotype as well as ASD clinical symptom scores in humans (Fig. 1L, 2A to H).

Next to assess the contribution of copper-deficiency in the pathogenesis of ASD, we took advantage of a copper-deficient mouse model treated with a copper (II) selective chelator, triethylenetetramine dihydrochloride (TETA) (55 mM) to eliminate internal copper (*24*) (fig. S2). Similar to the drug-induced ASD model mice, we administrated the TETA to the pregnant mother from embryonic days (E) 11.5 until the birth (Fig. 2I), and analyzed whether the reductions of copper levels in the mother and embryos. Consequently, we found that significant reductions of the blood and cortical levels of copper in TETA-treated mother and embryos at E18.5, respectively (Fig. 2J and K), confirming the validity of the TETA-treated mouse model as a copper-deficient model. We also assessed whether the chelating effect of TETA affects similar periodic elements, but no difference in cortical zinc concentrations was observed in TETA-treated embryos at E18.5 (data not shown), indicating that TETA-treated offspring serve as a selective model for copper-deficiency reflecting ASD pathophysiology. Compared with CTL, TETA-treated embryos showed small bodies (Fig. 2L) and reduced body weight (Fig. 2M). Moreover, TETA-treated embryos showed small brains (Fig. 2N) and reduced brain weight (Fig. 2O). These findings demonstrate that embryonic TETA treatment induces copper-deficiency, resulting developmental abnormalities in offspring.

### Copper-deficient offspring displayed ASD-relevant behaviors

To characterize the associations of copper-deficient offspring and ASD-like phenotypes, we analyzed mouse social behaviors and RRBs in TETA offspring mice at 7 weeks old (Fig. 3A). In three-chamber social interaction test (Fig. 3B), CTL mice preferred a novel mouse than an inanimate empty cage, and spent more time and distance when interacting with a social target during the social interaction trial (Fig. 3B, C to E). In contrast, TETA offspring mice spent similar time and distance when interacting with both a novel mouse and an inanimate empty cage (Fig. 3B, F to H). In the social novelty trial, CTL mice preferred a novel mouse than a familiar mouse, and spent more time, distance, and entry interacting with a novel mouse (Fig. 3B, I to K). TETA offspring mice also preferred a novel mouse than a familiar mouse, and spent more time, distance, but not in entry (Fig. 3B, L to N). Overall, these results indicate that TETA offspring mice show impaired social behaviors.

**Fig. 3.**
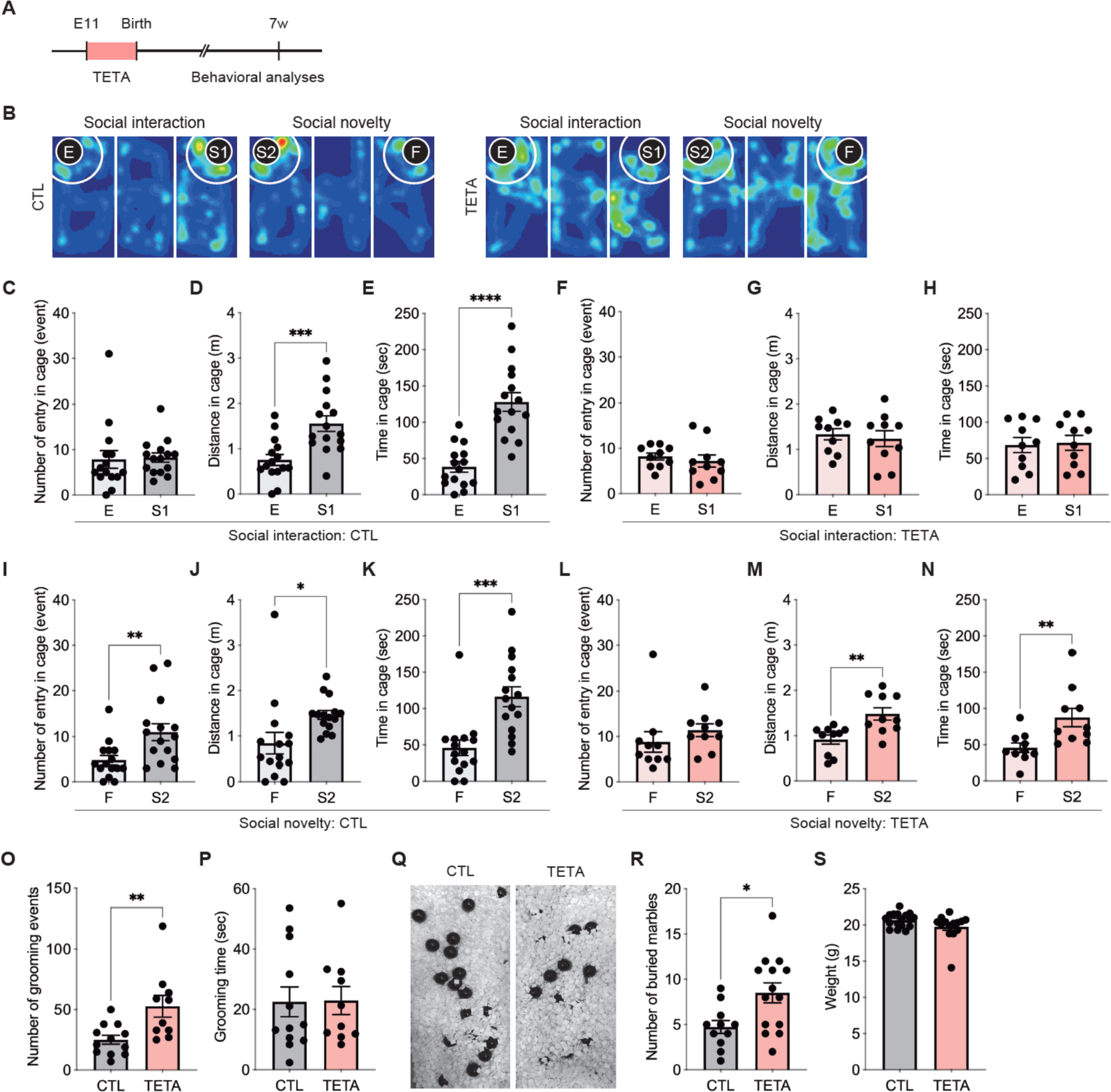
Copper-deficient offspring mice display ASD-relevant behaviors. (**A**) Experimental time course for TETA treatment to the pregnant mice for behavioral analyses. (**B**) Representative heatmaps of social interaction and social novelty behaviors in the three-chamber social interaction test at 7 weeks old. *E: empty, S1: stranger mouse one, S2: stranger mouse two, F: familiar mouse.* (**C** to **H**) Quantifications of entry numbers into the targeted cage area (C and F), distance traveled around the cage (D and G), and interaction time around the cage in white circles (E and H) in social interaction session in CTL (C to E) and TETA (F to H) mice, respectively. CTL mice socially interacted with a stranger mouse, but not in TETA mice. (I to N) Quantifications of entry numbers (I and L), distance (J and M), and interaction time (K and N) in social novelty session in CTL (I to K) and TETA (L to N) mice, respectively. Social novelty behavior was observed in TETA mice, but fewer than that in CTL mice. (**O** and **P**) Quantification of the number of grooming events (O) and grooming times (P) at 7 weeks old. (**Q**) Representative images after a marble-burying test at 7 weeks old. (**R**) Quantification of the number of buried marbles. TETA mice exhibited an increase in repetitive behaviors. (**S**) No difference in weight at 7 weeks old. Data are represented as means (±SEM). Asterisks indicate ****P < 0.0001, ***P < 0.001, **P < 0.01, *P < 0.05, unpaired t-test. n = 10-15/condition for three-chamber social interaction, n = 10-12/condition for repetitive behaviors, n = 11-14/condition for marble-burying tests, n = 14-17/condition for weight.

We next assessed RRBs in TETA offspring mice. We found that the number of grooming events was significantly increased in TETA offspring mice (Fig. 3O), but not in grooming time (Fig. 3P). In addition, we found that TETA offspring mice buried more marbles than CTL mice (Fig. 3Q and R). There was no difference in the weight at 7 weeks old (Fig. 3S). These results indicate that TETA offspring mice exhibit RRBs.

We further assessed other mouse behaviors such as locomotion activity, anxiety-like behavior, and impulsivity using open field and elevated plus maze tests. However, there were no behavioral differences in both open field (fig. S3A to H) and elevated plus maze tests (fig. S3I to P). Together, these results demonstrate that TETA-treatment in mice has selective effects on behavioral characteristics relevant to ASD.

### Copper-relevant transcriptome was associated with neurodevelopment

To gain insights into the molecular mechanisms underlying ASD-relevant behavioral phenotypes in TETA offspring mice, we performed the bulk RNA-sequencing (RNA-seq) of the prefrontal cortex (PFC) of TETA offspring mice at 7 weeks old. Transcriptomic profiles were distinctly separated between CTL and TETA offspring mice at 7 weeks old (Fig. 4A). Differential expression analysis of RNA-seq data identified 968 differentially expressed genes (DEGs) such as *Rxrg*, *Rgs9*, and *Adora2a* (false discovery rate [FDR] < 0.05) in the PFC of TETA offspring mice compared to CTL mice (Fig. 4B and C, table S2). We also conducted gene ontology (GO) analysis to infer the functional pathways of DEGs. Copper-relevant DEGs were involved in actin binding, metal ion transmembrane transporter activity, protein serine/threonine kinase activity, and transcription coregulator activity, and transmembrane transporter activity (Fig. 4D to F, table S3). These results indicate that transcriptome alterations in the PFC of TETA offspring mice impact on neurodevelopment and brain functions caused by copper-deficiency.

**Fig. 4.**
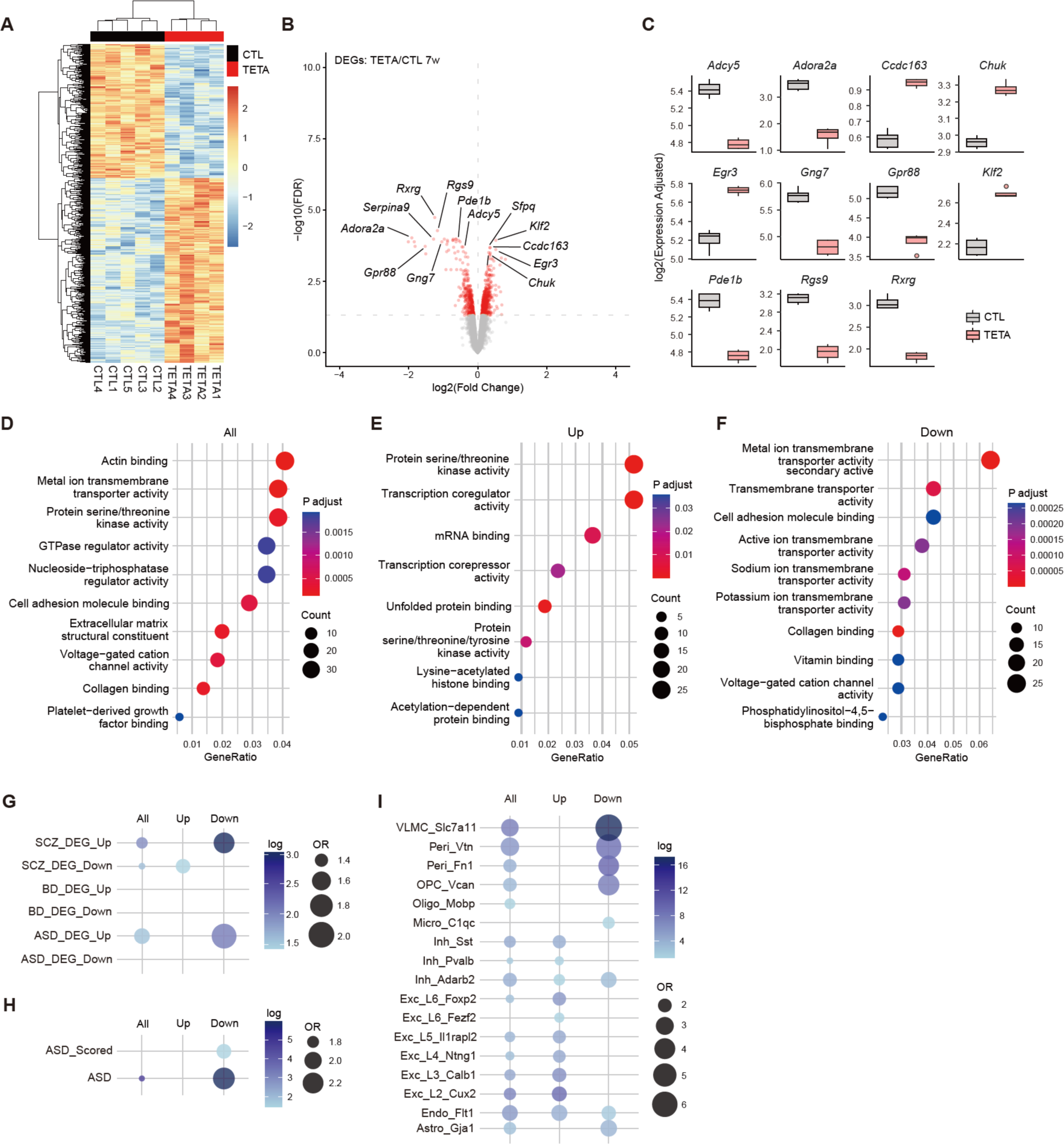
Transcriptomic alterations in copper-deficient offspring mice. (**A**) Heatmap of differentially expressed genes (DEGs) in the PFC of TETA offspring at 7 weeks old. (**B**) Volcano plot shows copper-relevant DEGs with the top 13 gene names. False discovery rate (FDR) < 0.05, Y-axis = -log10(FDR), X-axis = log2(Fold Change). (**C**) Bar plots of the top 11 genes of copper-relevant DEGs. (**D** to **F**) Gene ontology (GO) analyses of copper-relevant DEGs in biological process (BP). Scatterplots represent the top 10 functions in each module. Colors represent the adjusted p-values, dot sizes represent gene counts. *All: all DEGs* (D)*, Up: upregulated DEGs* (E)*, Down: downregulated DEGs* (F). (**G**) Dot plots of copper-relevant DEGs show the enrichment for disorder-specific human transcriptome (see Methods). *SCZ: schizophrenia, BD: bipolar disorder, ASD: autism spectrum disorder*. Dot plots represent the odds ratio (OR). Colors represent the - log10(FDR). (**H**) Dot plots of copper-relevant DEGs show the enrichment for SFARI ASD genes. Dot plots represent OR. Colors represent the -log10(FDR). *ASD Scored: scored SFARI ASD genes.* (**I**) Dot plots of copper-relevant DEGs show the cell-type specific enrichments. Dot plots represent OR. Colors represent the -log10(FDR). *VLMC: vascular leptomeningeal cells, Peri: pericytes, OPC: oligodendrocyte precursor cells, Oligo: oligodendrocytes, Micro: microglia, Inh: inhibitory neurons, Exc: excitatory neurons, Endo: endothelial cells, Astro: astrocytes*.

We also investigated the transcriptome during embryonic brain development using the PFC of TETA-treated embryos at E18.5 by bulk RNA-seq. Similar to the 7 weeks, we found clear separation of transcriptomic profiles between CTL and TETA-treated embryos (fig. S4A). We identified 442 DEGs such as *Ddit4*, *P4ha1*, and *Nid2* (FDR < 0.05) at E18.5 (fig. S4B and C, table S4). Functional analysis of embryonic DEGs identified pathways involved in nucleotide metabolic process, ribose phosphate metabolic process, purine nucleotide metabolic process, cellular component disassembly, and organic hydroxy compound biosynthetic process (fig. S4D and E, table S5). These results also indicate that metabolic abnormalities in the PFC of TETA-treated embryos at E18.5 are caused by copper-deficiency, suggesting that neurodevelopmental abnormalities are caused by copper-deficiency.

To further characterize 7 weeks old copper-relevant DEGs, we performed enrichment analyses for genes involved in psychiatric disorders and NDDs (see Methods). We found that downregulated copper-relevant DEGs were highly enriched in SCZ- and ASD-specific upregulated DEGs (Fig. 4G). Upregulated copper-relevant DEGs were also enriched in SCZ-specific downregulated DEGs (Fig. 4G). As we expected, downregulated copper-relevant DEGs were highly enriched in both scored and non-scored SFARI ASD genes (Fig. 4H). We also found that downregulated copper-relevant DEGs were enriched in SCZ-specific upregulated DEGs at E18.5 (fig. S5A and B). These results indicate that copper-relevant DEGs are affected in neurodevelopmental impairments such as SCZ and ASD.

### Oligodendrocytes-related genes were downregulated in copper-deficient offspring mice

To further identify what cell-type are affected by copper-deficiency, we next conducted cell-type enrichment analysis using single cell RNA-seq (scRNA-seq) datasets (see Methods) of the PFC compared with 7 weeks old copper-relevant DEGs. We found that the downregulated copper-relevant DEGs were highly enriched in vascular leptomeningeal cells (VLMCs), pericytes, and oligodendrocyte progenitor cells (OPCs) (Fig. 4I). Enrichments were also observed in microglia, inhibitory neurons, endothelial cells, and astrocytes (Fig. 4I). In contrast, the upregulated copper-relevant DEGs were enriched in inhibitory and excitatory neurons as well as endothelial cells (Fig. 4I). We also found that these trends in the copper-relevant DEGs at E18.5 (fig. S5C and D). Moreover, copper-relevant DEGs at E18.5 and 7 weeks old were both enriched in oligodendrocyte-lineage (fig. S6). These results demonstrate that copper-deficiency particularly impacts on the development of vascular cells and oligodendrocytes.

Among these cell-types, we focused on oligodendrocytes, because some of the most significant downregulated DEGs such as *Rxrg* and *Adcy5* were implicated in oligodendrocyte development (Fig. 4B and C, table S2). In addition, missense mutations of *ADCY5* have been identified in ASD, and *Adcy5* knockout mice display ASD-like behaviors including social impairment, RRB, and cognitive dysfunction (*25*). Thus, we examined whether the numbers of RXRG-positive (+) and ADCY5+ cells were reduced in the PFC. We found that significant reductions of the numbers of RXRG+ and ADCY5+ cells in the PFC of TETA offspring mice at 7 weeks old (Fig. 5A to D), underscoring a causal link between copper-deficiency and oligodendrocyte development.

**Fig. 5.**
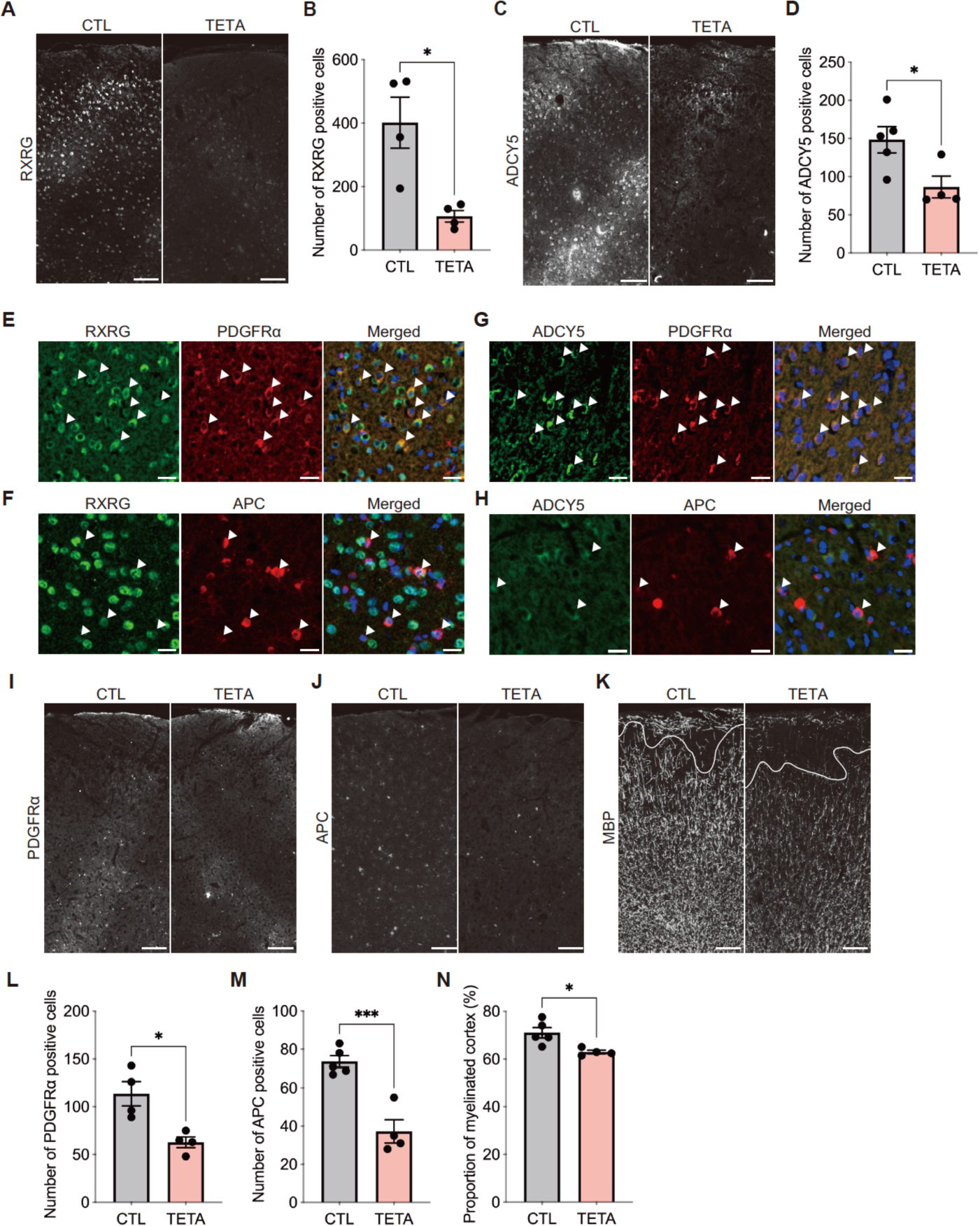
Impairments of oligodendrocyte development in copper-deficient offspring mice. (**A**) Representative fluorescent images of RXRG-positive (+) cells in the PFC at 7 weeks old. (**B**) Quantifications of RXRG+ cells in the PFC. (**C**) Representative fluorescent images of ADCY5+ cells in the PFC at 7 weeks old. (**D**) Quantifications of ADCY5+ cells in the PFC. Significant reductions of RXRG+ and ADCY+ cells were observed in the PFC of TETA mice. (**E** and **F**) Representative fluorescent images of RXRG expression in oligodendrocyte-lineage cells in the PFC at 7 weeks old. RXRG+ cells expressed both PDGFRα (E) and APC (F), markers for OPC and mature oligodendrocyte respectively. (**G** and **H**) Representative fluorescent images of ADCY expression in oligodendrocyte-lineage cells in the PFC at 7 weeks old. ADCY+ cells expressed both PDGFRα (C) and APC (D), markers for OPC and mature oligodendrocyte respectively. (**I** and **J**) Representative fluorescent images of PDGFRα+ or APC+ oligodendrocytes in the PFC at 7 weeks old. (**K**) Representative fluorescent images of myelinated cortex by myelin basic protein (MBP) immunostaining at 7 weeks old. (**L** and **M**) Quantifications of oligodendrocytes in the PFC. Significant reductions of PDGFRα+ OPC (L) and APC+ mature oligodendrocytes (M) were observed in the PFC of TETA mice. (**N**) Quantification of the proportion of myelinated PFC. A reduction of myelinated cortical area was observed in the PFC of TETA mice. Data are represented as means (±SEM). Asterisks indicate ***P < 0.001, *P < 0.05, unpaired t-test. n = 4-5/condition. Scale bars: 50 μm in A, B, and I to K, 20 μm in E to H.

### Copper-deficient offspring mice exhibited developmental defects in oligodendrocytes

Next, we investigated whether RXRG and ADCY5 were expressed in oligodendrocyte lineage cells. We found that RXRG and ADCY5 were both expressed in PDGFRα+ OPCs (Fig. 5E and G) and APC+ mature oligodendrocytes (Fig. 5F and H) in the mouse PFC. Moreover, we found reductions of the numbers of PDGFRα+ OPCs and APC+ mature oligodendrocytes were found in the PFC of TETA offspring mice (Fig. 5I, J, L and M). In addition, we examined the cortical myelination in the PFC of TETA mice due to loss of oligodendrocytes. The proportion of cortical myelinated area by immunostaining with myelin basic protein (MBP), a structural component of myelin, was significantly reduced in the PFC of TETA offspring mice (Fig. 5K and N). These results further underscore the key role of copper-deficiency in the development and maturation of oligodendrocytes in the PFC.

### Oligodendrogenesis via mTOR signaling was dysregulated by copper-deficiency

To explore the molecular mechanisms underlying oligodendrocyte defects, we reinvestigated the DEGs involved in oligodendrogenesis, and found that the several DEGs involved in mTOR signaling at both E18.5 and 7 weeks old, such as *Ddit4*, *Rasd2*, *Rxrg*, and *Adcy5* (Fig. 4B and C, fig. S4B and C). mTOR signaling is pivotal for oligodendrocyte development (*26, 27*).

Therefore, we evaluated whether mTOR signaling was dysregulated in the cortex of TETA offspring at protein level (Fig. 6A). We assessed the mTOR signaling by confirming the phosphorylation levels of downstream targets (Fig. 6B). We found that there was no difference in the expressions of RPS6 and 4EBP1 in the cortex of TETA-treated embryos at E18.5 (Fig. 6C, F, and H). Nevertheless, we interestingly found that the decreased phosphorylation levels of RPS6 and 4EBP1 in the cortex of TETA-treated embryos at E18.5 (Fig. 6C, G, and I). These results thus indicate that mTOR signaling is dysregulated in the cortex of TETA-treated embryos at E18.5.

**Fig. 6.**
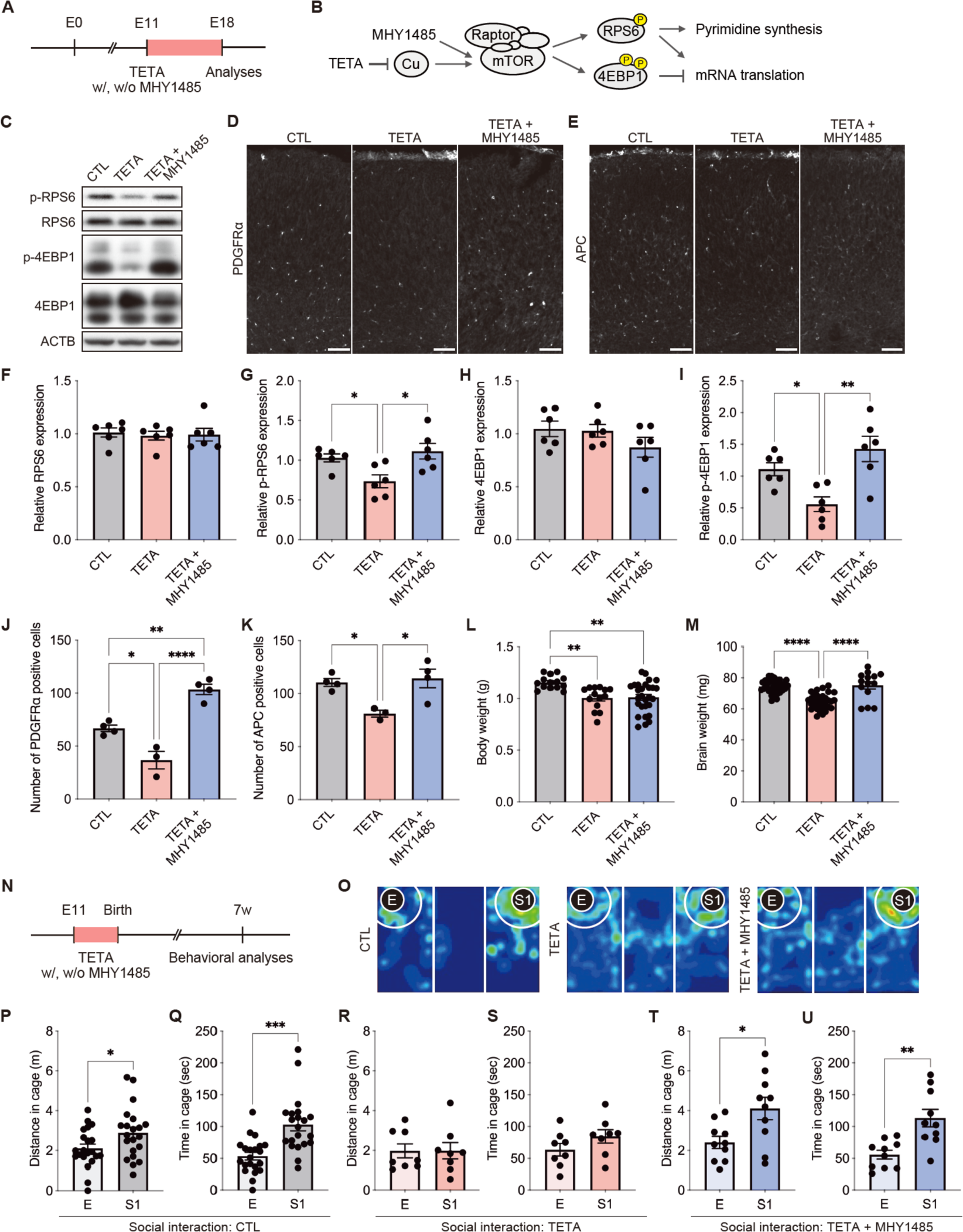
Activation of mTOR signaling rescues the phenotypes observed in copper-deficient offspring mice. (**A**) Experimental time course for TETA treatment with/without MHY1485 to the pregnant mice. (**B**) Simplified diagram of copper and mTOR signaling. (**C**) Representative immunoblotting of mTOR signal proteins in the cortex at E18.5. *CTL: CTL + saline, TETA: TETA + saline, MHY1485: TETA + MHY1485.* (**D** and **E**) Representative fluorescent images of PDGFRα+ (D) or APC+ (E) oligodendrocytes in the PFC at E18.5. (**F** to **I**) Quantification of RPS6 (F), phosphorylated (p)-RPS6 (G), 4EBP1 (H), and p-4EBP1 (I). Significant reductions of p-RPS6 and p-4EBP1 in TETA-treated embryos were rescued with an mTOR activator MHY1485 treatment. (**J** and **K**) Quantifications of oligodendrocytes in the PFC at E18.5. Significant reductions of PDGFRα+ (J) and APC+ (K) oligodendrocytes in TETA-treated embryos were rescued with MHY1485 treatment. (**L** and **M**) Quantifications of body weight (L) and brain weight (M) at E18.5. A significant reduction of brain weight in TETA-treated embryos was rescued with MHY1485, but not in body weight. (**N**) Experimental time course for TETA treatment with/without MHY1485 to the pregnant mice for behavioral analyses. (**O**) Representative heatmaps of social interaction behaviors in the three-chamber social interaction test at 7 weeks old. *E: empty, S1: stranger mouse one.* (**P** to **U**) Quantifications of distance traveled around the cage (P, R, and T) and interaction time around the cage in white circle (Q, S, and U) in social interaction session in CTL (P and Q), TETA (R and S), and TETA with MHY1485 (T and U) treated mice, respectively. MHY1485 treatment rescued social impairments in TETA mice. Data are represented as means (±SEM). Asterisks indicate ****P < 0.0001, ***P < 0.001, **P < 0.01, *P < 0.05, one-way ANOVA with a Tukey’s multiple comparison test, unpaired t-test. n=6/condition for Western blotting, n = 3-4/condition for cell counts, n = 14-28/condition for body weight, n = 14-34/condition for brain weight, n = 8-21/condition for behavioral tests at 7 weeks old. Scale bars: 50 μm in D and E.

We further investigated whether the dysregulation of mTOR signaling by copper-deficiency can be rescued with an mTOR activator MHY1485 (Fig. 6B). MHY1485 (10 mg/kg) was administered intraperitoneally every other day (at E11.5, 13.5, 15.5, and E17.5) during TETA treatment (Fig. 6A). In TETA-treated embryos with MHY1485 treatment, we found that there was no difference in the expressions of RPS6 and 4EBP1 in the cortex at E18.5 (Fig. 6C, F, and H). In addition, we found that the improved phosphorylation levels of RPS6 and 4EBP1 in the cortex of TETA-treated embryos with MHY1485 treatment at E18.5 (Fig. 6 C, G, and I). These results also indicate that the dysregulation of mTOR signaling in the cortex by copper-deficiency at E18.5 is rescued with mTOR activator.

Next, we investigated whether oligodendrogenesis could be rescued by the activation of mTOR signaling with MHY1485. As well as oligodendrocytes phenotype in TETA offspring at 7 weeks old, significant reductions of the numbers of PDGFRα+ OPCs and APC+ mature oligodendrocytes were also observed in the cortex of TETA-treated embryos at E18.5 (Fig. 6D, E, J, and K). Remarkably, we found that the numbers of PDGFRα+ OPCs and APC+ mature oligodendrocytes were rescued in the cortex of TETA-treated embryos with MHY1485 treatment at E18.5 (Fig. 6 D, E, J, and K). In addition, we found that reduced brain weight of TETA-treated embryos at E18.5 was also recovered with MHY1485 treatment (Fig. 6M), but not in body weight (Fig. 6L). Together, these results further demonstrate that the dysregulation of mTOR signaling by copper-deficiency results in impairments of oligodendrocyte development.

We further investigated whether ASD-relevant behaviors in TETA offspring mice could be rescued by oligodendrocytes via activation of mTOR signaling. At 7 weeks old, we first evaluated social behavior in MHY1485-treated TETA mice (Fig. 6N), and surprisingly found that reduced social interactions in TETA offspring mice was improved with MHY1485 treatment, compared with CTL and TETA offspring mice (Fig. 6O, P to U). We also assessed the effects of MHY1485 treatment on RRBs, however there were no effects on RRBs (fig. S7A to C). Regarding weight, there were no difference among group at 7 weeks old (fig. S7D). These results indicate that social impairment caused by copper-deficiency can be rescued by activation of mTOR signaling. These results also suggest that the association between oligodendrocytes and social behaviors in the pathogenesis of ASD.

### White matter volume was associated with ASD social symptoms

Finally, we analyzed T1-weighted MR images from individuals with ASD to investigate the impact of oligodendrocytes on social behavior in human. We found the decreased total volumes of white matter in individuals with ASD compared to CTL (Fig. 7A and B). Consistent with the phenotypes of copper-deficient offspring mice, MR imaging results suggest that developmental abnormalities in oligodendrocytes and myelination formation individuals with ASD. In addition, the volumes of white matter negatively correlated with ADOS-2 scores such as social interaction, social affect, and total scores, except for communication and RRB scores (Fig. 7C to H). These results demonstrate that white matter volumes are associated with social symptoms in individuals with ASD, suggesting oligodendrocytes impact on social function in human.

**Fig. 7.**
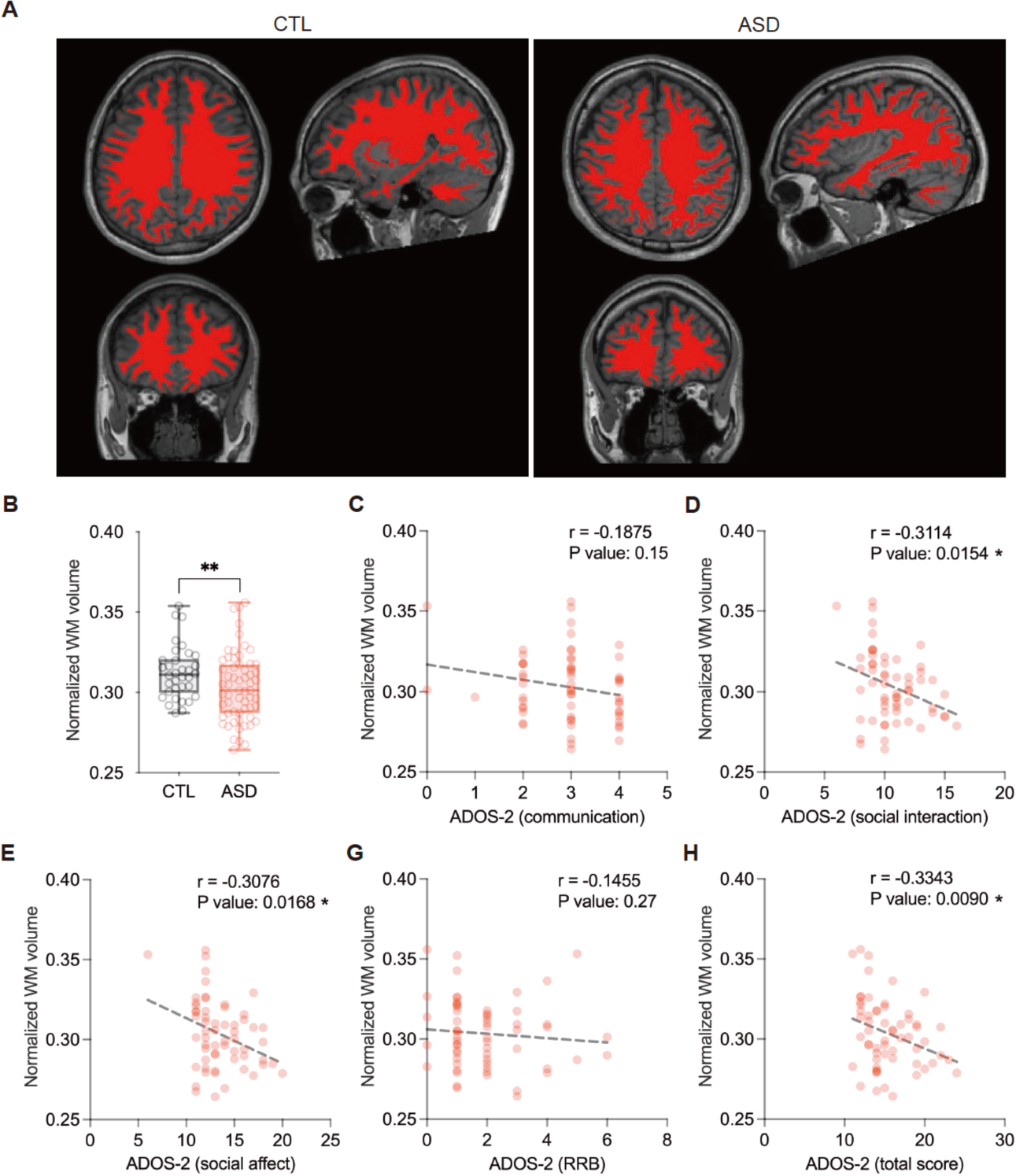
White matter volume correlates with social symptoms in individuals with ASD. (**A**) Representative T1-weighted images using 3-tesla magnetic resonance imaging. White matter (WM) region of interest was highlighted in red. (**B**) Quantifications of WM volume in CTL and individuals with ASD. Total volumes of WM were significantly decreased in individuals with ASD. (**C** to **H**) WM volume correlated with ADOS-2 scores. Total volumes of WM were negatively correlated with the social interaction score (D), social affect score (E), and total score (H) of ADOS-2 in individuals with ASD, but not with the communication score (C) and RRB score (G). Data are represented as box and whiskers (minimum to maximum). Asterisks indicate **P < 0.01, *P < 0.05, Spearman rank correlation coefficient test, multivariate linear regression analysis with sex, age, and hemolysis as confounders. n = 34-60/condition for total volume of WM, n = 60 for correlations between total volume of WM and ADOS-2 scores.

## DISCUSSION

In this study, we found dramatic alterations of metallome in individuals with ASD and negative correlations between copper and ASD symptoms. Copper-deficient mice displayed ASD-like behaviors and impairments of oligodendrogenesis and myelination. Copper-relevant DEGs were enriched in ASD. Moreover, we showed downregulation of mTOR signals in copper-deficient mice, and rescued social behavior and oligodendrocytes phenotypes by mTOR activation. Supporting these results, decreased white matter volumes in individuals with ASD showed negative correlation with social symptoms. Our study demonstrates that copper-deficiency is associated with oligodendrocyte impairments and ASD pathophysiology via mTOR dysfunction.

Copper is an essential element that accumulates in the brain and plays roles in cell survival and differentiation, synaptic plasticity, sleep and wakefulness, metabolism, redox, and immunity (*18, 28–32*). Menkes disease is an X-chromosome recessive genetic disorder characterized by copper-deficiency caused by mutations in copper transporter *ATP7A* showing severe central nervous system impairment, developmental delay, seizures, and hypothermia (*31, 32*). Interestingly, the expressions of myelin-associated genes, including *MBP* and *PLP1 (myelin proteolipid protein),* were downregulated in the postmortem brain of a patient with Menkes disease (*33*). Wilson’s disease is a disorder in which mutations in the copper transporter *ATP7B* cause copper to accumulate in the body, resulting in motor and cognitive dysfunction, decreased motivation, and other psychiatric symptoms (*31, 32*). However, in ASD, no conclusion has been reached about the increase or decrease of copper, and previous studies have used different samples such as whole blood, serum, hair, and saliva, as well as the races, regions, and measurement methods of the subjects, and there have been various reports of increases, no differences, or decreases (*18, 21, 34–45*). Additionally, the altered zinc-copper metabolic rhythmicity precedes the emergence of ASD (*46*).

Focusing on copper in blood samples from ASD individuals, an increased copper have been reported in serum of Chinese children with ASD (*34*), in plasma of autistic, Asperger’s and PDD-NOS Individuals from the USA (*37*), in serum of autistic Individuals from Venezuela (*38*), or in plasma of ASD Individuals from China (*21*), in plasma of Autistic Children from India (*39*), in serum of ASD children from Russia (*40*), on the other hand, there are reports of no significant changes of copper in serum of children with ASD in Slovenia (*41*), serum of ASD children from the USA (*42*), whole blood of ASD children from Romania (*43*), in serum of ASD children from North America (*44*). Remarkably, consistent with our results, there was a report showing decreased copper in serum of ASD children from Chinese Han Population using about 100 samples in each group (*45*). In summary, many studies report increased blood copper in ASD, whereas others report changes or decreases in blood copper. In the ASD animal studies, valproic acid-exposed female offspring rats showed a reduction of serum copper, but not in male rats (*35*). We also found that reduced serum copper in MIA offspring mice showed a positive correlation with mouse vocal communications (Fig. 2F, G, and H). These results support our human metallome results in individuals with ASD.

Copper-deficiency also affects redox by reducing functions of superoxide dismutase (SOD), as copper-containing SOD converts O_2_ to H_2_O_2_ under conditions of oxidative stress (*19*). Oxidative stress, lipid peroxidation, and mitochondrial dysfunction are involved in ASD pathogenesis (*47*). We have also reported that increases of oxidative stress-related metabolites in the plasma of children with ASD (*10*). Thus, copper-deficiency may affect redox and be associated with a worsening of the oxidative state in the pathogenesis of ASD. Moreover, specificity protein 1 (SP1), a zinc-finger transcription factor, functions by binding zinc, but in the presence of copper it binds with high affinity copper and regulate different gene expression (*48*). Increased expression of SP1 has been reported in the anterior cingulate gyrus of the postmortem brain in individuals with ASD, and SP1 regulates the downstream expressions of ASD genes, such as OXTR and PTEN (*49*). Zinc-binding proteins including transcription factors are encoded in 10% of human genome. These studies suggest that copper-deficiency causes functional alterations in transcription factors that adversely affect the regulation of ASD genes and genes required for brain development.

Together, these findings suggest that copper-deficiency contributes to the pathogenesis of ASD. Copper plays an essential role in various biological events, and its deficiency adversely affects brain development. Therefore, these observations underscore the significance of maintaining optimal levels of copper.

In this study, we identified oligodendrocyte-related genes in copper-relevant DEGs. In downregulated DEGs, RXRG and its associated gene ADCY5 play roles in maturation and myelination of oligodendrocytes as well as Schwann cells via mTORC1-RXRG-SREBP-lipid biosynthesis (*50–52*). These studies demonstrate that mTOR-RXRG signals regulate oligodendrogenesis and myelination. Indeed, mTOR signals control oligodendrogenesis during development mainly depending on mTORC1, which regulates myelin-associated lipogenesis and protein gene regulation (*26, 52, 53*). It is also known that cuprizone, a copper chelator, is used to make demyelinating disease model animals and induces metabolic dysfunction in oligodendrocytes resulted in inflammation, oxidative stress, cell death, and demyelination (*54*). Therefore, our results demonstrate that copper plays essential roles in oligodendrogenesis and myelination via mTOR-RXRG signals.

In addition, copper-relevant DEGs were also enriched in pericyte and endothelial cells (Fig. 4I), suggesting that these vascular cells may be impaired by copper-deficiency. Interestingly, pericytes are proliferated during demyelination to stimulate OPC differentiation and myelination (*55*). Pericytes also modulate blood-axon barrier, structure, function, and connectivity in white matter (*56*). Endothelial cells regulate OPC specification and migration during CNS development (*57, 58*). Endothelial cells also secrete trophic factors to support OPC survival and proliferation (*59*). These studies suggest that pericyte and endothelial cells crosstalk with oligodendrocytes to promote differentiation, migration, and myelination as well as maintaining white matter structure.

Myelination is regulated by mTOR (*52*), and white matter abnormalities have been reported in ASD (*60, 61*). The decreased volumes of white matter in individuals with ASD was found in this study (Fig. 7). In general, ASD findings in the tuberous sclerosis complex and improvement of social behaviors with rapamycin support the involvement of mTOR hyperactivity in ASD pathogenesis (*62–66*). In contrast, decreased mTOR signaling in postmortem brain of ASD has actually been reported (*67*). Consistent with our results, the studies have also reported that mTOR dysregulation in various ASD animal models (*67–69*). Indeed, balanced mTOR activity in oligodendrocytes is crucial for proper myelination, emphasizing the importance of maintaining an optimal level, neither too high nor too low (*53*). Together, these findings suggest that appropriate balance of mTOR is critical in ASD pathogenesis and white matter development.

Alterations of 14 trace elements (lithium, magnesium, vanadium, chromium, manganese, iron, cobalt, copper, zinc, selenium, tin, barium, mercury, and lead) have been reported in ASD (*18–23*). In this study, we further demonstrate the alterations of 5 novel elements in plasma of individuals with ASD: sodium, phosphorus, sulfur, and calcium as decreased elements, and potassium as an increased element (Fig. 1). Among these novel elements, we found that plasma concentrations of sodium and phosphorus showed correlations with social symptoms of ADOS-2 scores (data not shown). Several trace elements that have already been reported such as lithium, zinc, selenium, and barium were also showed correlations with ADOS-2 scores (data not shown). These results suggest that not only specific elements, but also comprehensive metallome contributes to the pathophysiology of ASD. However, the roles of these elements in ASD pathogenesis are unknown, and future studies using human samples and animal models are needed.

We acknowledge several limitations in this study. Although the sample size for the metallomics analysis was >100 each, large-scale studies with more participants are needed because ASD is heterogeneous. Although our study consisted only of Japanese, the genetic background was more homogeneous, so we conclude that the results are not confounded by race or diet, and reflect the general pathophysiology in individuals with ASD. However, study design and potential residual confounders including biometric information such as height and weight, socioeconomic factors, family history of neurodevelopmental or psychiatric disorders, and self-selection bias should be acknowledged. Additionally, CTL subjects were recruited through advertisement, thus there were possibilities that backgrounds and educational backgrounds vary.

In summary, we found the decreased plasma copper in ASD showed negative correlations with ASD symptoms. We also found that reduced mTOR signaling in a copper-deficient model mice cause oligodendrocyte developmental abnormalities, resulting in social impairment. Consistent with these findings, decreased white matter volumes in individuals with ASD showed negative correlations with social symptoms. Our study suggests that metallome dyshomeostasis links to the pathophysiology of ASD through dysfunction of biological events involved in trace elements. To our knowledge, this is the first study to demonstrate the biological mechanisms of copper-deficiency in ASD using animal models. Our findings provide the novel insights into the metallome dyshomeostasis in understanding biological mechanisms underlying ASD.

## MATERIALS AND METHODS

### Study design

The overall objective of this study was to understand the biological mechanisms underlying ASD by focusing on the metallome. A case-control study was conducted to evaluate the plasma metallome in Japanese individuals with ASD, following STROBE guidelines (*70*). To identify the important trace element, the correlations between metallome and clinical symptoms were evaluated. All procedures for clinical studies were approved by the University Ethics Committees and were conducted in accordance with the Ethical Guidelines for Medical and Health Research Involving Human Subjects of the Ministry of Health, Labour and Welfare of Japan. All participants were fully informed about the study before enrollment, and their parents and/or legal guardians provided written informed consent. Minimum samples size was determined using G*Power 3.1.9.6 (*71, 72*). Participant enrollments and demographics were summarized in fig. S1 and table S1, respectively. To investigate the biological mechanism of trace element, a mouse model was generated, and the phenotypes were analyzed. All procedures for animal experiments were approved by the University Animal Research Committee and were performed in accordance with ARRIVE guidelines. No statistical methods were used to predetermine the sample size for animal studies, which was determined empirically. Lastly, human brain MR imaging data was analyzed to investigate the findings from the mouse model. Detailed Materials and Methods for all experiments were described in Supplementary Materials. The experimenters were blinded to genotype allocation during experiments and statistics.

### Statistical analysis

Data are represented as means of biological independent experiments with box and whiskers (minimum to maximum) or ±standard error of the mean (SEM). Statistical analyses (D’Agostino-Pearson test, multivariate linear regression analysis, unpaired *t*-test (two-sided), Mann-Whitney test (two-sided), Pearson’s r correlation coefficient, Spearman rank correlation coefficient test, linear regression, one-way ANOVA with a Tukey’s multiple comparison test) were performed using Prism 9 (GraphPad Software, Boston, MA, USA) and SPSS 29.0.1 (IBM, Armonk, NY, USA). The D’Agostino-Pearson test was used to examine normal distribution. Multivariate linear regression analysis with sex, age, and hemolysis as confounders was used for metallome analysis. Unpaired *t*-test and Mann-Whitney test were used for demographic characteristics and animal studies. Spearman rank correlation coefficient test was used to examine the correlations between trace elements and ADOS-2 scores. Linear regression was used to calculate the line that best fit the correlation analysis. Discriminant analysis was performed to calculate the discrimination score to distinguish the CTL and ASD groups. The numbers of samples and used statistics were described in each corresponding figure legend. Asterisks indicate p-values (****P < 0.0001, ***P < 0.001, **P < 0.01, *P < 0.05). P < 0.05 was considered to indicate statistical significance.

## Data Availability

The data supporting the findings of this study are available from the corresponding author upon reasonable request.

## Acknowledgments

We are grateful to Dr. Genevieve Konopka for beneficial discussion. We also thank Seiichi Inagaki, Katsura Kato, Naoyuki Okamoto, Takuya Shimizu, RENATECH Co., Ltd. for metallomics analysis supports.

## Funding

This work was founded by

The Japan Agency for Medical Research and Development (AMED) Translational Research Grant Seeds A140 (NU, SS)

AMED-CREST 22gm1510009h0001 (MM)

AMED-PRIM 21wm04250XXs0101 (MM)

AMED 21uk1024002s0201 (MM), 20gm6310015h0001 (MM)

The Japan Society for the Promotion of Science (JSPS) Grant-in-Aid for Scientific Research (B) 23H02837 (NU), 19H03581 (HM), 20H03604 (MM)

JSPS Grant-in-Aid for Scientific Research (C) 20K06872 (NU)

JSPS Grant-in-Aid for Early-Career Scientists 23K14443 (MD)

JSPS Grant-in-Aid for Challenging Research (Exploratory) 19K21754 (HM)

Uehara Memorial Foundation (NU)

Takeda Science Foundation (NU) Naito Foundation (NU)

Mochida Memorial Foundation for Medical and Pharmaceutical Research (NU)

Inamori Foundation (NU)

SENSHIN Medical Research Foundation (NU)

Osaka Medical Research Foundation for Intractable Diseases (NU, MD)

MEI Grant supported by Osaka University (NU)

Osaka University Medical Doctor Scientist Training Program (NN)

## Author contributions

Conceptualization: NU

Methodology: NU, MD, SB, KM

Software: SB

Validation: NU, MD, SB, KM, KI, NN, HM, YF, TY, TK, HM, MM, SS

Investigation: NU, MD, SB, KM, RI, KI, NN, HM, YF, TY, TH, MT, MJX, YK, NU, KI, KO, TH, TY, TK, MT, HM, MM

Formal analysis: NU, MD, SB, KM

Resources: NU, SB, KM, RI, TY, TH, MT, MJX, YK, NU, KI, KO, TH, MT, HM, MM

Data Curation: SB, KM Writing - Original Draft: NU

Writing - Review & Editing: NU, MD, SB, HM, MM, SS

Visualization: NU, MD, SB, KM, KI

Supervision: NU, HM, MM, SS

Project administration: NU

Funding acquisition: NU, MD, HM, MM, SS

## Data and materials availability

The data supporting the findings of this study are available from the corresponding author upon reasonable request. The NCBI Gene Expression Omnibus (GEO) accession number for the RNA-seq data are available at GSE232864. Custom R codes and data to support the analysis, visualizations, functional, and gene set enrichments are available at https://github.com/BertoLabMUSC/Usui_Metallome_2023.

## Supplementary Materials

### Materials and Methods

#### Ethics statement

All procedures were approved by the Ethics Committee of Osaka University (#19394), University of Fukui (#20200112), and Nara Medical University (#1319), and were conducted in accordance with the Ethical Guidelines for Medical and Health Research Involving Human Subjects of the Ministry of Health, Labour and Welfare of Japan. All participants were fully informed about the study before enrollment, and their parents and/or legal guardians provided written informed consent.

#### Participants

A case-control study was conducted to evaluate the metallome in Japanese individuals with ASD following STROBE guidelines (*73*). Individuals with ASD were recruited through advocacy groups in cooperation with Asperger Society Japan (Nagoya, Japan), University of Fukui (Fukui, Japan) and Nara Medical University (Nara, Japan). CTL participants were also recruited in Fukui and Nara, Japan by advertisement. According to the pilot study, the minimum number of required samples was 100 participants for each group using G*Power 3.1.9.6 (two-tails, effect size = 0.4, significance level = 0.05, power = 0.80)(*74, 75*). Initially, 270 eligible individuals were identified as individuals with ASD and CTL. ASD has been diagnosed by >2 experienced psychiatrists based on the criteria outlined in the Diagnostic and Statistical Manual of Mental Disorders, Fifth Edition (DSM-5) following clinical interviews, the Japanese version of ADOS-2, and the Wechsler Adult Intelligence Scale-IV (WAIS-IV) FIQ ≥ 70. CTL participants were assessed using the WAIS-IV FIQ ≥ 70, and the Autism-Spectrum Quotient (AQ-J) < 26. Instead of AQ-J, the Autism Spectrum Screening Questionnaire (ASSQ) < 18 was used in the selection of children aged <18 years. The Mini-International Neuropsychiatric Interview (MINI) or Structured Clinical Interview (SCID) was also employed to assess for any personal or family history of past or present mental illness for CTL participants. All participants were interviewed by a physician for a comprehensive assessment of their medical history to exclude those with neurological or medical impairments. Specifically, patients with fragile X syndrome, epileptic seizures, obsessive–compulsive disorder, affective disorders, schizophrenia, additional psychiatric or neurological diagnoses were excluded. Participants with organic causes such as tumors on magnetic resonance imaging were excluded. All participants were required to be drug naive including any dietary supplements for at least 6 months before this study. Of the 155 individuals with ASD, 13 were excluded because of consent withdrawal; 21, diagnosed not based on the DSM5 criteria; 3, medications; 5, extreme vagal response; 1, intellectual disability (FIQ < 70); thus 112 individuals were registered with ASD (Extended Data Fig. 1). Of the 115 CTL participants, none had withdrawn consent were excluded, thus 115 CTL participants were recruited (Extended Data Fig. 1). Eventually, 112 ASD and 115 CTL participants were enrolled (Extended Data Fig. 1, Supplementary Tables S1); however significant differences were found between the groups in terms of sex and age, which were considered confounders. The characteristics of the participants are summarized in Supplementary Table S1.

#### Human blood sampling

Blood sampling was performed as previously described (*76*). Fasting blood samples were collected by venepuncture in a sitting position with a tourniquet between 7:00 and 15:00 hours. Plasma samples were immediately centrifuged at 1000*g* for 10 min, then supernatants were collected in aliquots of 200-400 µL and stored at −80°C.

#### Metallomics analysis

Metallomics analysis was performed as previously described (*77*). All instruments used for trace element measurements, including sample storage bins, sampling bins, and micropipette tips, were made of polypropylene. All containers were washed with ultrapure water (≥18.00 M Ω•CM or more), added 500 μL of 60% nitric solution (#64003-03, Kanto Chemical Co., Ltd., Tokyo, Japan), and then heated overnight. The containers were washed again, filled with 10% nitric acid, and heated for 2 overnights. For the pretreatment of blood samples, 50 μL of blood sample, 125 μL of 60% nitric acid solution, and 25 μL of 30% hydrogen peroxide solution were mixed in a pluggable polypropylene container and heated at 70°C for 16 hours. For the pretreatment of tissue samples, tissue samples were weighed and acidly digested using a microwave digestion system (ETHOS EASY, Milestone Srl, Sorisole BG, Italy) with a container in a closed pressure vessel system. According to the digestion program (temperature was raised to 180°C in 20 min and held at 180°C for 10 min), 925 μL of 60% nitric acid and 25 μL of 30% hydrogen peroxide water were added to the container. After acid degradation, the sample solutions were filled to 10 mL with ultrapure water. A standard solution (#XSTC-622B, SPEX, Metuchen, NJ, USA) and a thallium standard solution (#Tl-1000, Kanto Chemical Co., Japan) were mixed for the calibration curve, then appropriately diluted with 3% nitric acid solution, and calibration curves of lithium, manganese, iron, cobalt, copper, zinc, arsenic, selenium, rubidium, strontium, molybdenum, silver, cesium, barium, and thallium were prepared. In addition, single element standard solutions were mixed and appropriately diluted with 3% nitric acid solution to create calibration curves of sodium, magnesium, phosphorus, sulfur, potassium, and calcium. Correlation coefficients were greater than 0.9998 for all 21 elements. Equipment conditions were high frequency output of 1550 W, plasma gas flow rate of 15 L/min, nebulizer gas flow rate of 1.05 L/min, and auxiliary gas flow rate of 0.9 L/min. Samples were introduced into the apparatus by aspiration using a peristaltic pump. Under the above conditions, 21 elements including lithium (ppb), sodium (ppm), magnesium (ppm), phosphorus (ppm), sulfur (ppm), potassium (ppm), calcium (ppm), magnesium (ppb), iron (ppb), cobalt (ppb), copper (ppb), zinc (ppb), arsenic (ppb), selenium (ppb), rubidium (ppb), strontium (ppb), molybdenum (ppb), silver (ppb), cesium (ppb), barium (ppb), and thallium (ppb) were quantified by Agilent 7800 ICP-MS (Agilent Technologies, Santa Clara, CA, USA).

#### Mice

All procedures were performed in accordance with ARRIVE guidelines. The animal study protocol was reviewed and approved by the Animal Research Committee of Osaka University (approval number #27-010). Timed-pregnant wild-type C57BL/6J mice (Japan SLC Inc., Shizuoka, Japan) were obtained by timed breeding. For embryo staging, the day of detection of vaginal plug was considered E0.5. Mice were housed in the barrier facilities of Osaka University under a 12 h light– dark cycle and had free access to water and food. All mice used for behavioral testing and weight measurement were 7 weeks old male. The experimenters blind to genotypes performed experiments. All behavioral tests were performed between 9:00 to 17:00 hours.

#### Poly(I:C) treatment

Poly(I:C) was administered as described previously (*78*). Poly(I:C) (20 mg/kg; #P9582, Merck, Darmstadt, Germany) was dissolved in saline (#3311401A2026, Otsuka Pharmaceutical Co., Ltd., Tokyo, Japan), and intraperitoneally injected at E12.5. Saline was used for CTL.

#### TETA treatment

Copper (II) selective chelator, triethylenetetramine dihydrochloride (TETA) (55 mM, #T5033, Merck) was dissolved in tap water, and administered to pregnant mice from E11.5 until birth by free drinking. Tap water was given on the same schedule for CTL.

#### Primary mouse cortical neuron culture

Neurons were dissected from E17.5 embryonic cortex in ice-cold HBSS (#H6648, Sigma-Aldrich, St. Louis, MO), digested with 0.1% papain (#P3125, Merck, Darmstadt, Germany) in MEMα (#135-15175, FUJIFILM Wako Pure Chemical Corporation, Osaka, Japan), and triturated with 60 mg/ml DNase I (#D4527, Merck) and 10% fetal bovine serum (FBS). Dissociated cells were plated at a density of 20,000 cells/well on poly-L-lysine-(#P2636, Merck) coated culture cover glasses (#C1100, Matsunami Glass, Osaka, Japan). Medium was replaced to Neurobasal (#21103049, Thermo Fisher Scientific, Waltham, MA, USA) supplemented with B27 (#17504044, Thermo Fisher Scientific) and 0.5 mM GlutaMAX (#35050061, Thermo Fisher Scientific) 3 hours after the neurons had adhered. Neurons were treated with copper (II) inhibitors; triethylenetetramine dihydrochloride (TETA) (#S6585, Selleck Chemicals, Houston, USA) or ammonium tetrathiomolybdate (TTM) (#013-26932, FUJIFILM Wako Pure Chemical Corporation) with/without copper (II) chloride dihydrate (#09505-72, Nacalai Tesque) at 3 days *in vitro* (DIV). Proliferating undifferentiated neurons and glia were eliminated at 3 DIV using cytosine β-D-arabinofuranoside hydrochloride (AraC) (#C1768, Merck).

#### Immunocytochemistry

Immunohistochemistry was performed as previously described (*79*). Cells were fixed with 4% PFA in PBS for 10 min at room temperature, placed in PBS, and then permeabilized in PBS-T. Blocking was performed using 10% goat serum and 1% BSA in PBS-T for 1 hour at room temperature. Cells were incubated with the following primary antibodies overnight at 4°C: guinea pig anti-MAP2 (1:1,000; #188004, Synaptic Systems, Goettingen, Germany) and mouse anti-Tau-1 (1:1000; #MAB3420, Merck). Cells were then washed and incubated with secondary antibodies for 1 hour at room temperature. For fluorescence immunostaining, species-specific antibodies conjugated to Alexa Fluor 488 and/or Alexa Fluor 594 (1:2,000; Thermo Fisher Scientific) were applied, and coverslips were mounted with Fluoromount/Plus (#K048, Diagnostic BioSystems). DAPI (#11034-56, Nacalai Tesque) was used to stain the nucleus. Images were collected using All-in-One fluorescence microscope (BZ-X700, KEYENCE Corporation).

#### Sholl analysis

Sholl analysis was performed as previously described (*80*). Morphological characteristics of neurons were quantified blindly to genotype. Concentric circles having 10 μm increments in radius were defined from the center of cell body. The number of MAP2-positive dendrites crossing each circle was counted.

#### Mouse blood sampling

Blood samples were collected from pregnant female mice at E18.5. The serum samples were kept at room temperature for 30 min and centrifuged at 1500*g* for 10 min, then supernatants were collected and stored at -80°C until use.

#### Mouse tissue sampling

Mouse embryonic brains were collected from embryos at E18.5. The brain samples were weighted and stored at -80°C until use.

#### MHY1485 treatment

An mTOR activator, MHY1485 (10 mg/kg; 4,6-Dimorpholino-N-(4-nitrophenyl)-1,3,5-triazin-2-amine, #M3107, Tokyo Chemical Industry Co., Ltd., Tokyo, Japan) was dissolved with dimethyl sulfoxide (#D2438, Merck) and Sesame Oil (#25620-65, Nacalai Tesque, Kyoto, Japan), and intraperitoneally injected every other day from E11.5 until birth. The dissolve solution without MHY1485 was used for CTL.

#### USVs

USVs analysis was performed as described previously (*78*). Pups were removed from the dam and placed in individual soundproof rooms. Recordings were taken for 3 min using an UltraSoundGate condenser microphone (CM16, Avisoft Bioacoustics, Glienicke/Nordbahn, Germany) placed at a fixed height of 20cm above the pups, and recorded with UltraSoundGate 416H hardware and Avisoft RECORDER software (Avisoft Bioacoustics; approximately 20 dB gain, sampled at 16 bits, 250 kHz). Acoustic spectrograms were created in MATLAB (50% overlap, 512-point Hamming windows), yielding a temporal resolution of 1.024 ms and a spectral resolution of 488.3 Hz. Spectrograms were bandpass filtered from 20 to 120 kHz and filtered for white noise. Data quantification and analysis included the number and duration of calls (ms), number of calls with and without frequency jumps, mean frequency (kHz), frequency range of calls (kHz), and mean call slope (Hz/ms).

#### Three-chamber social interaction test

Three-chamber social interaction test was performed as previously described (*81*). Social interaction test consisted of three 5 min trials in the three-chamber apparatus (W600 × D400 × H220 mm, SC-03M, Muromachi Kikai Co., Ltd.). In the first trial, the mouse was allowed to explore freely the three-chamber apparatus. Each end chamber contained an empty wire cage (φ90 × H185 mm), with the middle chamber empty. In the second 5 min trial, to examine the social interaction, a stranger was placed in a wire cage in one end chamber and an empty wire cage in the opposite end chamber. The third 5 min trial examined the social novelty by placing the same mouse used in the second trial in the same wire cage in one end chamber and a new, unknown mouse in a wire cage in the opposite end chamber. The test mouse was also allowed to choose between an inanimate cage and a novel stranger in the second trial and between a familiar mouse and a stranger in the third trial. Interaction with targets around the wire cage was measured using ANY-maze behavior tracking software (Stoelting Co., Wood Dale, IL, USA).

#### Stereotyped behaviors test

Stereotyped behaviors test was performed as previously described (*81*). Mice were placed in a novel home cage, where they were habituated for 10 min, followed by a 10 min recording period. Time spent and the number of grooming events were manually quantified from the recorded videos.

#### Marble-burying test

Marble-burying test was performed as previously described (*81*). Mice were placed in the corner of a novel home cage with 18 marbles evenly distributed and allowed to explore freely for 20 min. After 20 min, the number of buried marbles was recorded. A marble was defined as buried if less than one-third of the marbles were visible.

#### Open field test

Open field test was performed as previously described (*81*). Mice were placed in one of the corners of a novel chamber (W700 × D700 × H400 mm, #OF-36(M)SQ, Muromachi Kikai Co., Ltd., Tokyo, Japan) and allowed to explore freely for 10 min. Time spent in the center of the arena (140 × 140 mm) and all corners of the arena (140 × 140 mm × 4 corners), and locomotor were measured by ANY-maze behavior tracking software (Stoelting Co.).

#### Elevated plus maze test

Elevated plus maze test was performed as previously described (*81*). Mice were placed in the center of the maze (open arms W54 × D297 mm; closed arms W60 × D300 × H150 mm; height from floor 400 mm, #EPM-04M, Muromachi Kikai Co., Ltd.) and allowed to explore freely the maze for 5 min. Time and distance in each arm were measured by ANY-maze behavior tracking software (Stoelting Co.).

#### RNA-seq

RNA-seq was performed using a service by Macrogen Japan Corp. (Kyoto, Japan) as previously described (*81*). Briefly, total RNA was extracted from mouse PFC at E18.5 and 7 weeks old using the miRNeasy Mini Kit (#217004, Qiagen, Hilden, Germany). RNA integrity number (RIN) of total RNA was quantified by Agilent 2100 Bioanalyzer (Agilent Technologies) using Agilent RNA 6000 Pico Kit (#5067-1513, Agilent Technologies). Total RNA with RIN values of ≥8.7 were used for RNA-seq library preparation. mRNA was purified from 500 ng total RNA, and used for cDNA library preparation by TruSeq Stranded mRNA Library Prep (#20020594, Illumina, San Diego, CA, USA). The quality of cDNA Libraries were determined by 2100 Bioanalyzer using Agilent High Sensitivity DNA Kit (#5067-4626, Agilent Technologies). Libraries were sequenced as 101 bp paired-end on Illumina NovaSeq6000.

#### RNA-seq alignment and quality control

Reads were aligned to the mouse mm10 reference genome using STAR (v2.7.1a) (*82*). For each sample, a BAM file was created containing mapped and unmapped reads across splice junctions. Secondary alignments and multi-mapped reads were removed using in-house scripts. Only uniquely mapped reads were retained for further analyses. Quality control metrics were assessed by the Picard tool (http://broadinstitute.github.io/picard/). Gencode annotation for mm10 (version M25) was used for reference alignment annotation and downstream quantification. Gene level expression was calculated using featureCounts (v2.0.1) (*83*). Counts were calculated based on protein-coding genes from the annotation files.

#### Differential expression

Counts were normalized using counts per million reads (CPM). Genes without reads in either CTL or TETA samples were removed. To infer potential experimental confounders, surrogates variables were calculated using the *sva* package in R (*84*). Differential expression analysis was performed in R using DESeq2 (v1.34) (*85*) using Limma (v3.50) (*86*) with the following model: gene expression ∼ Treatment + nSVs. Log2 fold changes and p-values were estimated. P-values were adjusted for multiple comparisons using the Benjamini-Hochberg correction (FDR). Differentially expressed genes were defined as FDR<0.05. Mouse Gene IDs were converted to Human Gene IDs using *biomaRt* package in R (*87*).

#### GO analysis

Functional annotation of differentially expressed and co-expressed genes was performed using clusterProfiler (v4.2) (*88*). Benjamini-Hochberg FDR (FDR<0.05) was applied as a multiple comparison adjustment.

#### Cell-type specific enrichment

To identify cell-type specific enrichment of DEGs, scRNA-seq data sets from the mouse brain curated from PSY DEGS (*89*), SFARI ASD genes (https://gene.sfari.org), scMouse PFC (*90*), scMouse Young (CZ CellXGene) (*91*), and scMouse Olig databases were used (*92*). Fisher’s exact test in R with parameters: alternative = “greater”, conf.level = 0.99 were used. OR and Benjamini-Hochberg FDR were applied.

#### Immunohistochemistry

Immunohistochemistry was performed as previously described (*81*). Mouse brains at 7 weeks old were fixed overnight at 4°C in 4% PFA in phosphate-buffered saline (PBS), cryoprotected in 30% sucrose in PBS, embedded in Tissue-Tek O.C.T. Compound (#4583, Sakura Finetek Japan Co.,Ltd., Osaka, Japan) for cryosectioning. Cryosections (20 μm thick) were placed in PBS. Antigen retrieval pretreatment was performed by incubating the sections in citrate buffer (10 mM citrate, 0.05% Tween-20, pH 6) at 95°C for 10 min. Sections were stained with the following primary antibodies: rabbit polyclonal anti-ADCY5 (1:250; #BS3922R, Bioss Antibodies Inc., Woburn, MA, USA), rabbit polyclonal anti-RXRG (1:200; #18-360, ProSci Inc., Poway, CA, USA), rat monoclonal anti-PDGFRα (CD140a) (1:200; #14-1401-82, Thermo Fisher Scientific, Waltham, MA, USA), mouse monoclonal anti-APC (1:250; #OP80, Merck), and rabbit polyclonal anti-MBP (1:200; #ab40390, Abcam, Cambridge, UK). The sections were then washed and incubated with secondary antibodies for 1 hour at room temperature. For fluorescence immunostaining, species-specific antibodies conjugated to Alexa Fluor 488 and/or Alexa Fluor 594 (1:2,000; Thermo Fisher Scientific) were applied, and cover glasses mounted with Fluoromount/Plus (#K048; Diagnostic BioSystems, Pleasanton, CA, USA). DAPI (#11034-56; Nacalai Tesque) was used for nuclear staining. Images were collected using All-in-One fluorescence microscope (BZ-X700, KEYENCE Corporation, Osaka, Japan). Cell counts were quantified manually or using the Hybrid cell count application software (KEYENCE Corporation). The myelinated prefrontal cortex (PFC) area was quantified as described previously (*81, 93*).

#### Western blotting

Mouse brains were homogenized in lysis buffer (50 mM Tris-HCl, pH 7.5, 150 mM NaCl, 5 mM EDTA, 1% Triton X-100) containing protease inhibitors (1:100) (#P8340; Merck) using a Dounce homogenizer. Proteins of each sample were separated by SDS-PAGE, transferred to an Immun-Blot PVDF Membrane (#162-0177, Bio-Rad Laboratories, Hercules, CA, USA), blocked with blocking buffer (1% skim milk in Tris-buffered saline (TBS) with 0.1% Tween-20) for 30 min at room temperature, and incubated with the following primary antibodies overnight at 4°C: rabbit monoclonal anti-S6 ribosomal protein (1:1,000; #2217, Cell Signaling Technology, Danvers, MA, USA), rabbit monoclonal anti-phospho-S6 ribosomal protein (1:2,000; #5364, Cell Signaling Technology), rabbit monoclonal anti-4E-BP1 (1:1,000; #9644, Cell Signaling Technology), rabbit monoclonal anti-phosphor-4E-BP1 (1:1,000; #2855, Cell Signaling Technology), and rabbit polyclonal anti-beta actin (1:2,000, #ab8227, abcam). The membranes were washed with TBS-T (TBS with 0.1% Tween-20), blocked again, reacted with appropriate horseradish peroxidase (HRP)-linked species-specific IgG secondary antibodies (1:10,000; Cell Signaling Technology) for 1 h at room temperature. The membranes were washed and developed using SuperSignal West Pico Chemiluminescent Substrate (#PI-34080, Thermo Fisher Scientific). Images were collected using the Blue Basic Autorad Film (#F9023-8×10, BioExpress, Kaysville, UT, USA).

#### MRI

MRI data acquisition was performed as previously described using a 3-tesla clinical scanner equipped with a 32 phased-array head coil (Magnetom Verio; Siemens, Erlangen, Germany) (*94, 95*). The participants were scanned with a three-dimensional T1-weighted gradient echo sequence (repetition time [TR] = 1,900 ms; echo time [TE] = 2.54 ms; field of view [FOV] = 256 × 256 mm; acquisition matrix = 256 × 256; and 208 contiguous axial slices of 1 mm thickness). For voxel-based morphometry (VBM) analysis, MR images were aligned to the anterior-posterior commissure axis to set the origin to the anterior commissure and set the images parallel to the axis using the auto_reorient.m (http://www.nemotos.net/?p=17) on MATLAB (MathWorks, Natick, MA, USA). Individual T1-weighted MR images were segmented into the gray matter (GM), white matter (WM), and cerebrospinal fluid (CSF) volumes using the Statistical Parametric Mapping (SPM12) software package (Wellcome Trust Centre for Neuroimaging, London, UK) (http://www.fil.ion.ucl.ac.uk/spm). Then, total intracranial volume (ICV) was calculated as the sum of the GM, WM, and CSF volumes (*96*). In order to normalize for variation in the individuals’ head sizes, the WM volume was divided by each individual’s total intracranial volume (ICV).

---

**Fig. S1.**
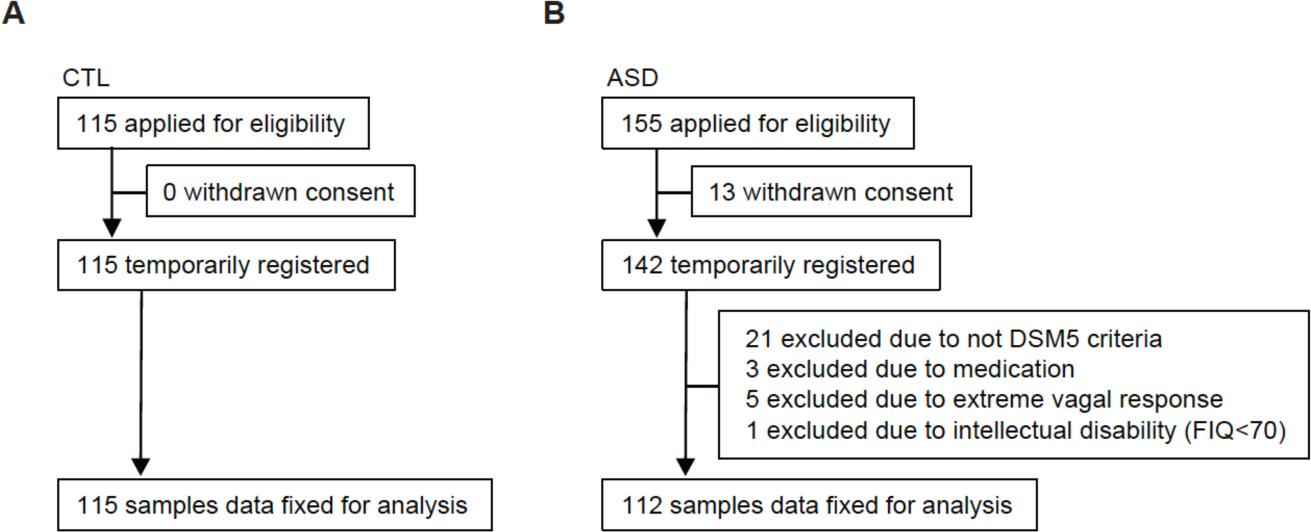
Flowcharts of trial participants for metallomics analysis. (**A** and **B**) Flowchart illustrating a case-control study using plasma of CTL (A) and ASD (B) individuals for metallomics profiling.

**Fig. S2.**
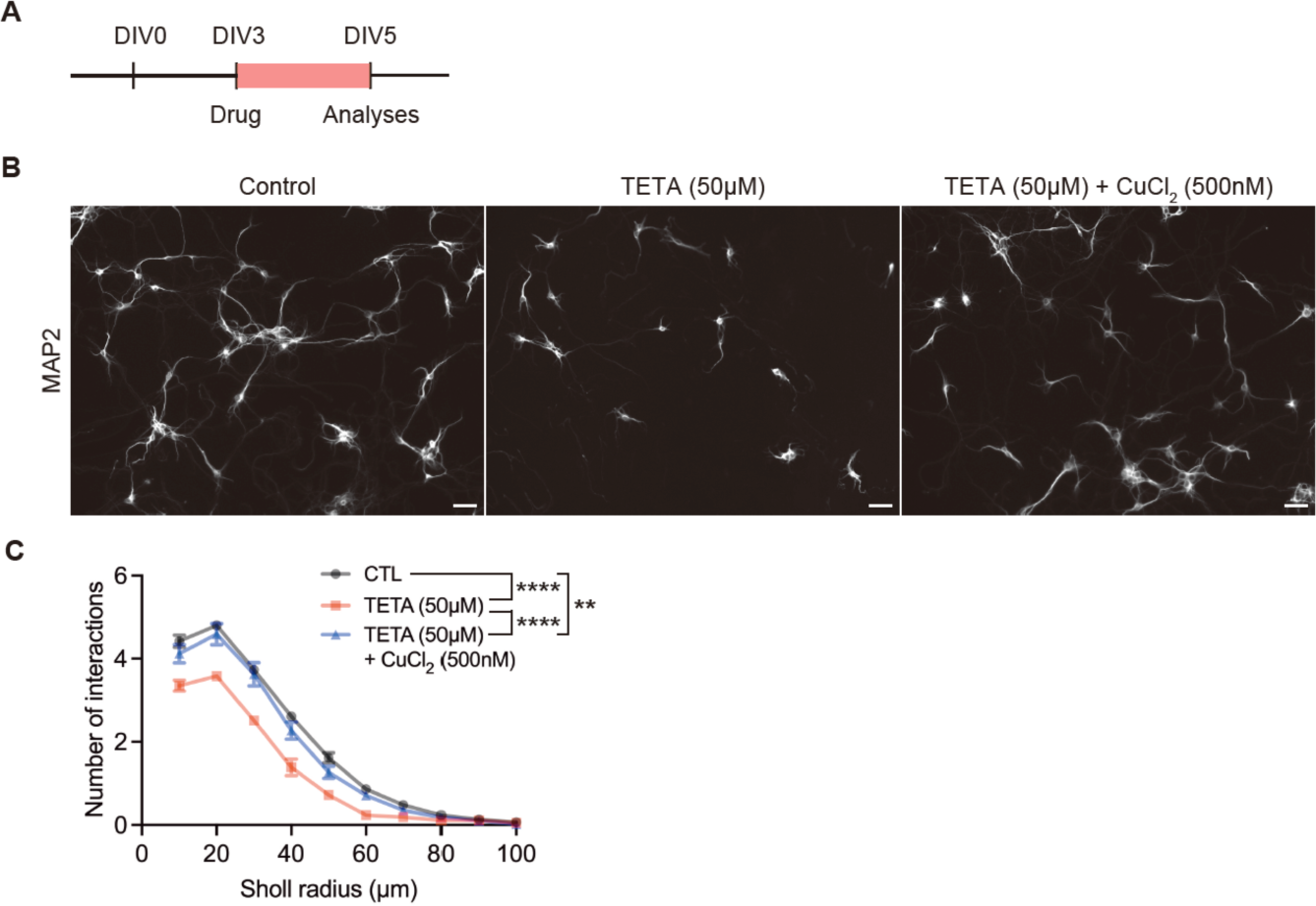
Triethylenetetramine dihydrochloride is a copper selective chelator. (**A**) Experimental time course for confirming the selectivity of triethylenetetramine dihydrochloride (TETA) against copper. (**B**) Copper is essential for dendritic formation of primary cultured neurons. (**C**) Quantification of dendritic formation in primary cultured neurons by Sholl analysis. The impaired formation of neuronal dendrites caused by copper deficiency in the presence of TETA was rescued by the administration of copper chloride (II). Data are represented as means (±SEM). Asterisks indicate ****p<0.0001, **p<0.01, two-way ANOVA with a Tukey’s multiple comparison test, n=97-185 cells/condition from 3 independent experiments. *TETA, triethylenetetramine*. Scale bars: 100 μm in B.

**Fig. S3.**
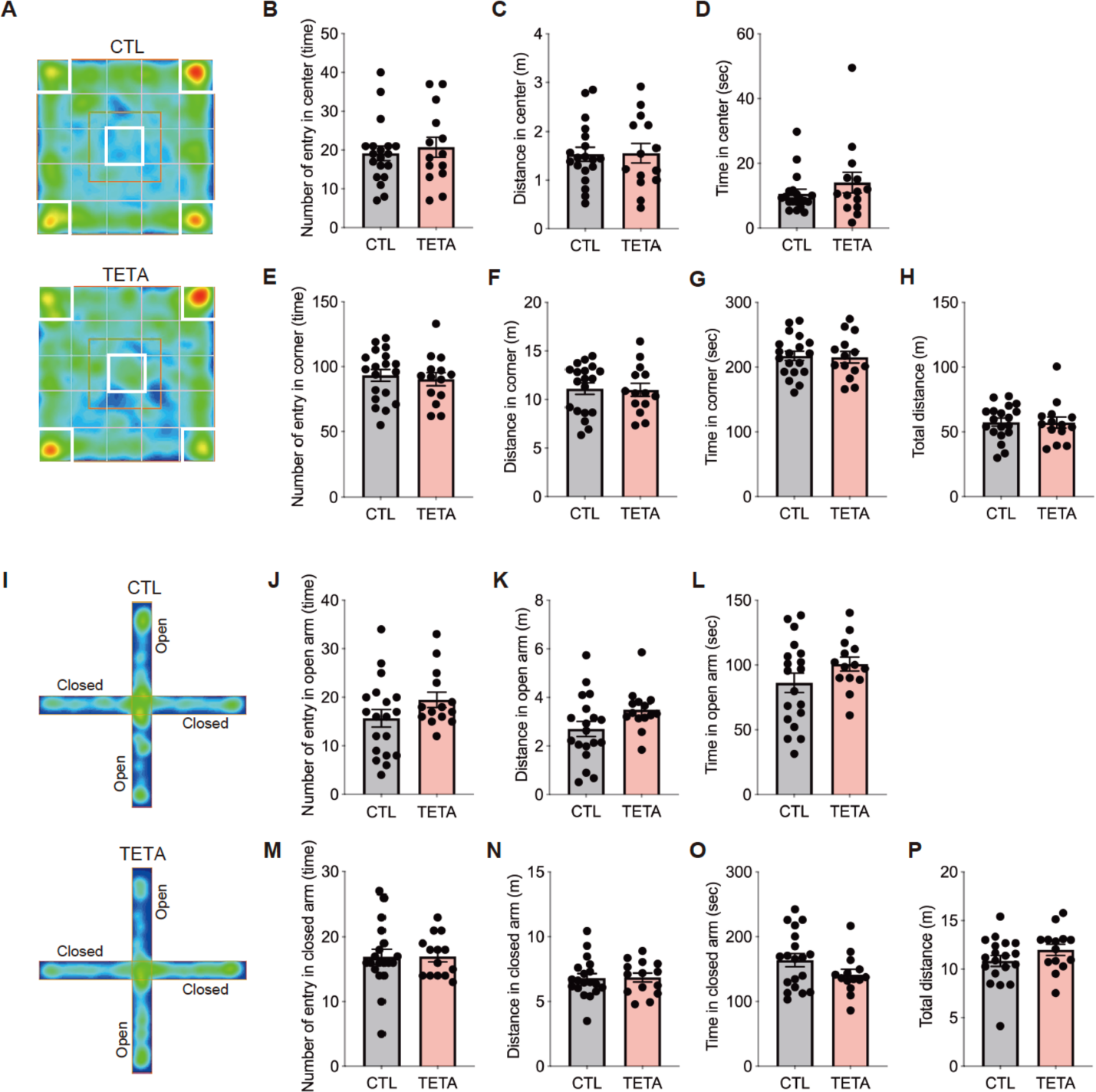
Copper-deficient offspring mice do not show anxiety-like behaviors. (**A**) Representative heatmaps of anxiety-like behavior in the open field test. (**B** to **H**) Quantification of the number of entries in center (B), distance in center (C), time in center (D), the number of entries in corner (E), distance in corner (F), time in corner (G), and total distance (H) during the open field test. (**I**) Representative heatmaps of anxiety-like behavior in the elevated plus maze test. (**J** to **P**) Quantification of entry numbers in open arm (J), distance in open arm (K), time in open arm (L), entry numbers in closed arm (M), distance in closed arm (N), time in closed arm (O), and total distance (P) during the elevated plus maze test. Data are represented as means (±SEM). Unpaired t-test. n = 14-19/condition.

**Fig. S4.**
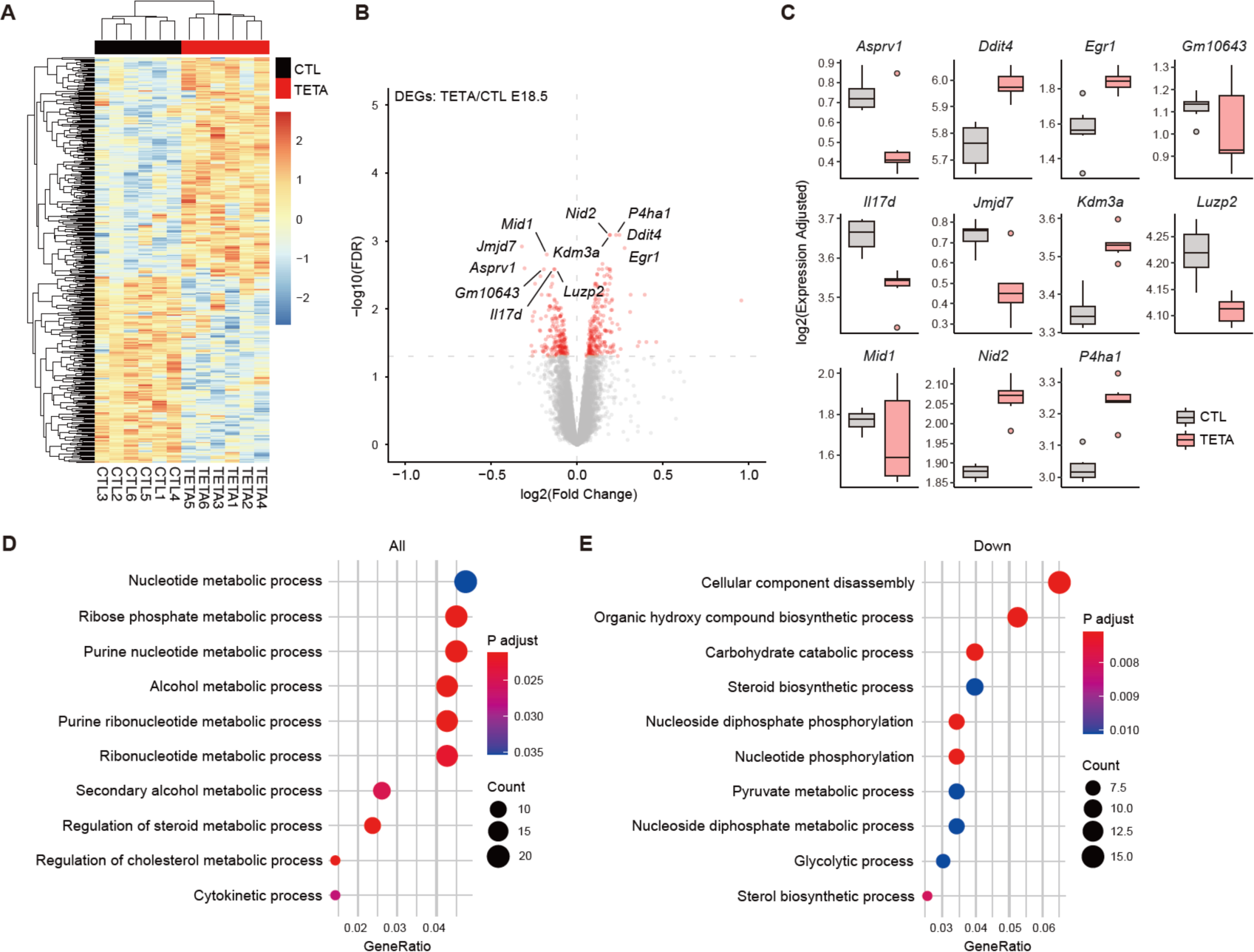
Transcriptomic alterations in copper-deficient embryos at E18.5. (**A**) Heatmap of differentially expressed genes (DEGs) in the PFC of TETA-treated embryos at E18.5. (**B**) Volcano plot shows copper-relevant DEGs with the top 11 gene names. FDR < 0.05, Y-axis = -log10(FDR), X-axis = log2(Fold Change). (**C**) Bar plots of the top 11 genes of copper-relevant DEGs. (**D** and **E**) GO analyses of copper-relevant DEGs in BP. Scatterplots represent the top 10 functions in each module. Colors represent the adjusted p-values, dot sizes represent gene counts. *All: all DEGs* (D)*, Down: downregulated DEGs* (E). There is no GOs in upregulated DEG in the PFC of TETA-treated embryos at E18.5.

**Fig. S5.**
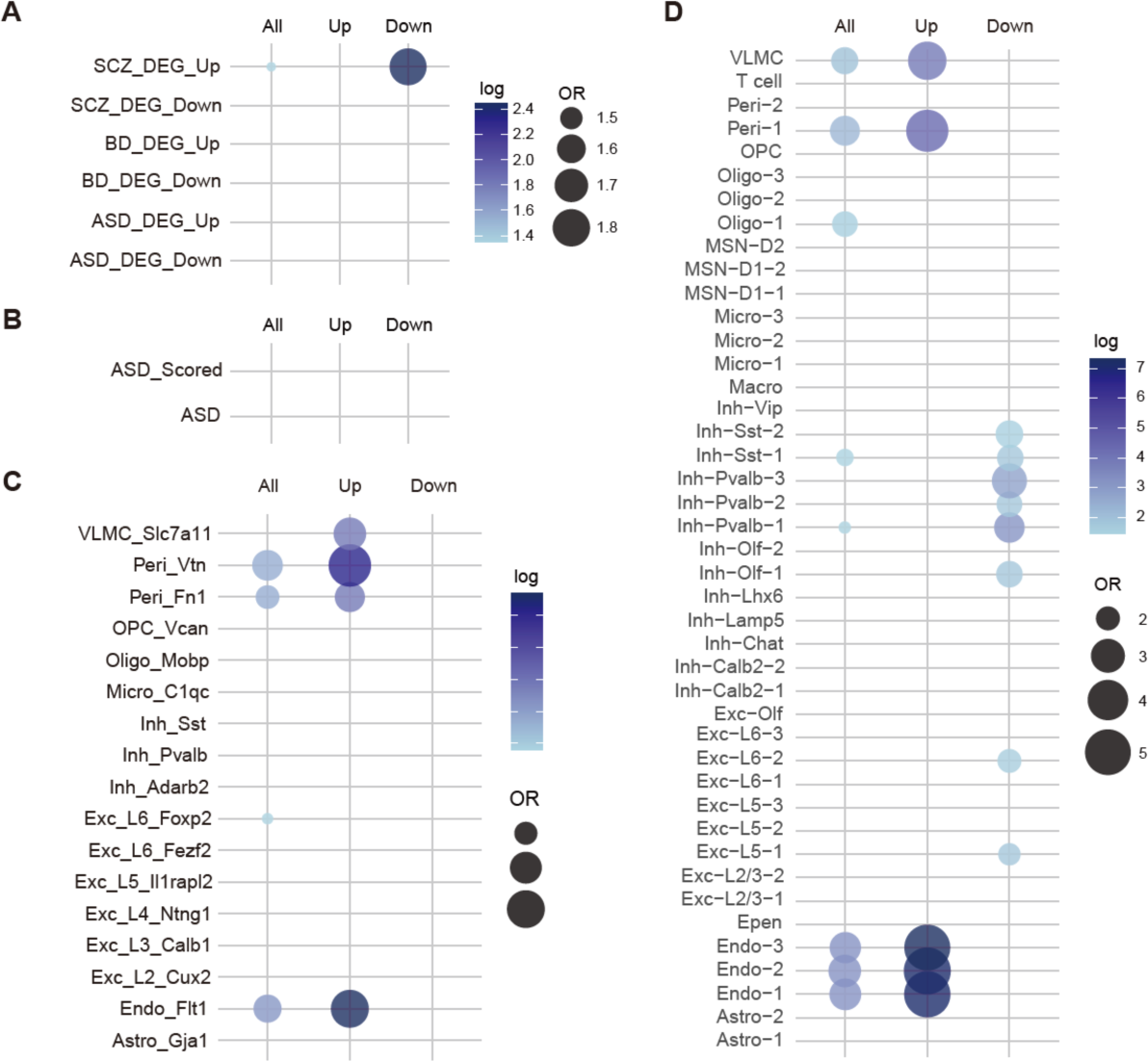
Enrichments of copper-relevant DEGs in disorder-specific transcriptome and brain cell-type at E18.5. (**A**) Dot plots of copper-relevant DEGs show the enrichment for disorder-specific human transcriptome (see Methods). *SCZ: schizophrenia, BD: bipolar disorder, ASD: autism spectrum disorder*. Dot plots represent OR. Colors represent the -log10(FDR). (**B**) No enrichment for SFARI ASD genes. *ASD Scored: scored SFARI ASD genes.* (**C**) Dot plots of copper-relevant DEGs show the cell-type specific enrichments. Dot plots represent OR. Colors represent the -log10(FDR). *VLMC: vascular leptomeningeal cells, Peri: pericytes, OPC: oligodendrocyte precursor cells, Oligo: oligodendrocytes, Micro: microglia, Inh: inhibitory neurons, Exc: excitatory neurons, Endo: endothelial cells, Astro: astrocytes*. (**D**) Dot plots of copper-relevant DEGs show the cell-type specific enrichments. Dot plots represent OR. Colors represent the -log10(FDR). *MSN: medium spiny neuron, Macro: macrophage, Epen: ependymal cells*.

**Fig. S6.**
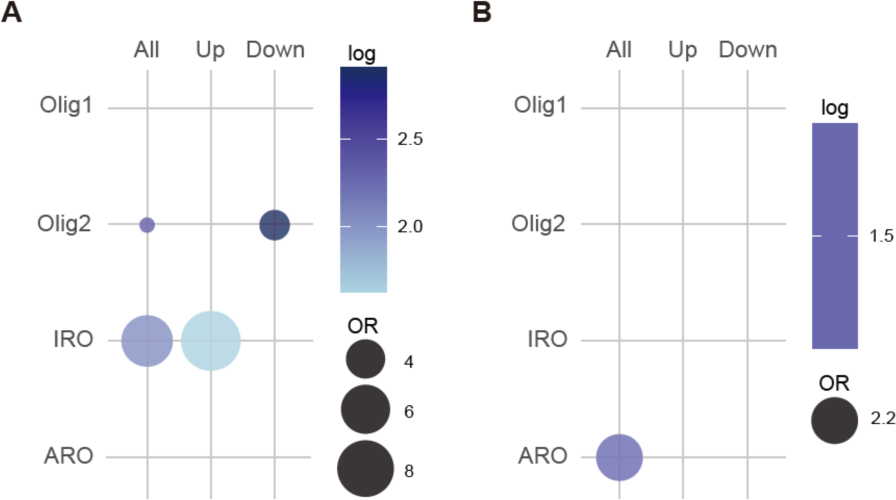
Oligodendrocyte-lineage enrichments of copper-relevant DEGs. (**A** and **B**) Dot plots of copper-deficient DEGs at E18.5 (A) and 7 weeks old (B) showing the enrichments in oligodendrocyte-lineage cells using scRNA-seq dataset (see Methods). Dot plots represent OR. Colors represent the -log10(FDR). *Olig1: Olig1-linage oligodendrocytes, Olig2: Olig2-linage oligodendrocytes, IRO: IFN-responsive oligodendrocytes subpopulation, ARO: age-related oligodendrocytes, All: all DEGs, Up: upregulated DEGs, Down: downregulated DEGs*.

**Fig. S7.**
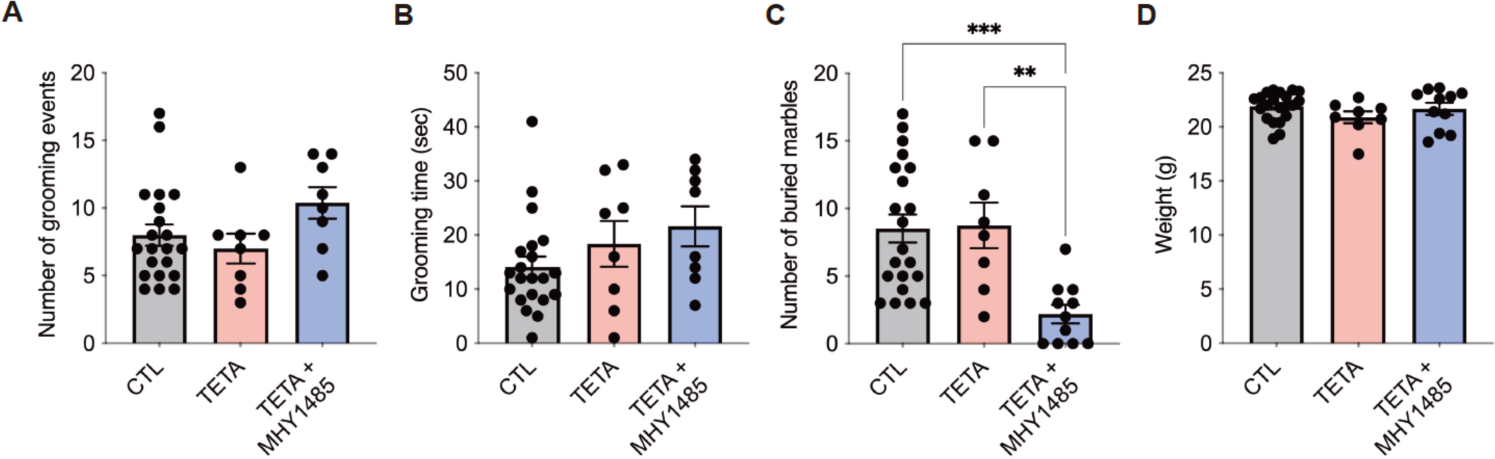
Activation of mTOR signaling did not improve repetitive behaviors. (**A** and **B**) Quantification of the number of grooming events (A) and grooming times (B) at 7 weeks old. (**C**) Quantification of the number of buried marbles. MHY1485 treatment could not improve repetitive behavioral phenotypes. (**D**) No difference in weight at 7 weeks old. Data are represented as means (±SEM). Asterisks indicate ***P<0.001, **P < 0.01, one-way ANOVA with a Tukey’s multiple comparison test, n = 8-21/condition for behavioral tests and weight at 7 weeks old.

